# Rapid and modular workflows for same-day sequencing-based detection of bloodstream infections and antimicrobial resistance determinants

**DOI:** 10.1101/2024.10.09.24315014

**Authors:** Mohammad Saiful Islam Sajib, Katarina Oravcova, Kirstyn Brunker, Paul Everest, Manuel Fuentes, Catherine Wilson, Michael E. Murphy, Taya Forde

**Affiliations:** School of Biodiversity, One Health & Veterinary Medicine, University of Glasgow, Glasgow, United Kingdom; MRC-University of Glasgow Centre for Virus Research, Glasgow, United Kingdom; Department of Microbiology, NHS Greater Glasgow and Clyde, Glasgow Royal Infirmary, New Lister Building, Alexandra Parade, Glasgow, UK; School of Medicine, Dentistry & Nursing, College of Medical, Veterinary & Life Sciences, Wolfson Medical School Building, University of Glasgow, Glasgow, UK

**Keywords:** rapid diagnosis, selective host depletion, chemical host depletion, metagenomic next generation sequencing, Oxford Nanopore technologies, selective sequencing, adaptive sampling, bloodstream infection, antimicrobial resistance

## Abstract

**Background:** Bloodstream infections (BSI) are a major global health concern, and existing diagnostic methods are too slow to guide targeted antibiotic therapy for critically ill patients, risking poor clinical outcomes. Rapid metagenomic-sequencing (mNGS) can facilitate swift pathogen and antimicrobial resistance (AMR) detection, but identification is challenged by significant host versus bacterial DNA in blood. To accelerate microbiological diagnosis, we developed M-15, a rapid and modular mNGS-based host DNA depletion workflow, validated with suspected BSI blood-culture samples and rapid culture-enriched spiked blood.

**Methods:** To assess chemical host DNA depletion (CHDD) efficiency, M-15 was benchmarked with five commercial/published protocols. Later, M-15 was combined with rapid mNGS with/without adaptive sampling (AS) and tested on clinical blood-culture samples (n=33) from suspected BSI cases identified on BACT/ALERT VIRTUO (30 flagged positive, three remained negative). To determine whether it is possible to utilise M-15 mNGS prior to blood-culture flagging positive, a rapid enrichment method was tested starting with 1-10 colony forming units of the top 15 bacterial species causing BSI spiked into BACTEC medium enriched with 10 mL sheep blood.

**Results:** All six chemical depletion protocols reduced host DNA by 2.5×10^0^ to 4.1×10^6^-fold, with the in-house M-15 protocol performing best, while adaptive sampling depleted host >5-fold. With BACT/ALERT specimens, M-15 mNGS accurately identified 3/3 negative, 28/28 mono-bacterial, and 2/4 multi-bacterial species. With rapid culture-enrichment and M-15 mNGS, <18% DNA was classified as host and all bacterial species tested (n=10) were correctly identified. M-15 mNGS accurately predicted phenotypic AMR/susceptibility for 90.3% (232/257) of drug/bacteria combinations from BACT/ALERT positive samples.

**Conclusions:** This study demonstrates that M-15 mNGS can facilitate species and AMR gene detection within 5-7 hours of BACT/ALERT positivity. Including 8-hour culture enrichment, microbiological and AMR confirmation is possible within 13-15 hours of sample collection. Thus, the M-15 mNGS workflow has the potential to improve patient outcomes in BSI.

## Background

Bloodstream infection (BSI) is one of the leading causes of death worldwide, with 2.9 million cases each year [1]. Rapid identification of the causative bacterial pathogens and their antimicrobial resistance (AMR) profiles is crucial for guiding more effective treatment and better patient outcomes, especially for critically ill patients such as those who develop sepsis [2]. Blood culture, the gold standard diagnostic method, often takes 48-72 hours, which is too slow to guide targeted antibiotic therapy. This necessitates the prescription of empirical antibiotics, which may fail to cover the resistance profile of the causative pathogen and risk poor clinical outcomes [3]. Also, the use of broad-spectrum antibiotics to limit disease progression may disrupt beneficial gut microbiota and contribute to the development of multidrug-resistant bacteria often associated with healthcare infections, further limiting the reserve of critically important antimicrobial agents [4]. Thus, there is an urgent need for rapid diagnostic tools to better inform antibiotic regimens at the early stages of BSI.

Certain applications of metagenomic next-generation sequencing (mNGS) could be used to rapidly identify bacterial species and AMR determinants, and therefore have the potential to accelerate BSI diagnosis [5]. However, the proportion of host DNA in blood compared to that of bacteria makes it challenging to directly sequence and generate adequate reads from the pathogen to accurately identify species and AMR in a cost-effective manner and within a clinically relevant timeframe. To address this issue, we developed a rapid mNGS-based chemical host DNA depletion (CHDD) protocol, named M-15, and compared its performance with two commercial kits (C1-2) and three published methods (P1-3) [6–8]. Next, M-15 mNGS was validated with and without nanopore adaptive sampling using both culture (BACT/ALERT an automated blood culture system) positive and negative enriched blood samples collected from patients with suspected BSI. Finally, M-15 was combined and tested with a rapid (8-hour) culture (BACT/ALERT) enrichment step to reduce turnaround time and enable same-day microbiological diagnosis of bloodstream infections.

## Methods

### Bacterial strains and host matrix

Defibrinated sheep blood (E&O Laboratories, UK) and EDTA-treated whole blood from healthy human volunteers were used as model host environment matrix. The list of clinical and/or American Type Culture Collection (ATCC) strains tested/used in this study can be found in **Tab. 1, S1, S2 and S3**.

**Tab. 1:** Host depletion efficiency and bacterial DNA loss observed with in-house chemical host DNA depletion method, M-15, compared with two commercial kits (C1-2) and 3 published protocols (P1-3).

| Organism | Methods/Kits | Ct (SD)<br>No chemical<br>depletion | Ct (SD)<br>Chemical depletion | ΔCt | Fold Reduction | DNA Removal (%) |
| --- | --- | --- | --- | --- | --- | --- |
| **Host depletion efficiency** |  |  |  |  |  |  |
| *Ovis aries* | M-15 (this study) | 16.24 (0.79) | 36.76 (1.82) | -20.5 | $4.1 \times 10^6$ | 99.99998% |
| *Ovis aries* | P1 [6] | 16.24 (0.79) | 33.69 (0.99) | -17.5 | $5.5 \times 10^5$ | 99.99982% |
| *Ovis aries* | Zymo HostZero (C1) | 20.30 (0.58) | 29.74 (2.10) | -9.43 | $5.9 \times 10^2$ | 99.82966% |
| *Ovis aries* | Molysis Complete 5 (C2) | 20.51 (0.64) | 25.73 (1.08) | -5.2 | $4.8 \times 10^1$ | 97.93324% |
| *Ovis aries* | P2 [7] | 18.97 (0.49) | 21.15 (0.53) | -2.2 | $5.3 \times 10^0$ | 81.09315% |
| *Ovis aries* | P3 [8] | 18.97 (0.49) | 20.15 (0.51) | -1.2 | $2.5 \times 10^0$ | 59.19500% |
| **Bacterial loss** |  |  |  |  |  |  |
| *E. coli* | M-15 (this study) | 23.69 (1.04) | 23.91 (0.47) | -0.22 | $1.3 \times 10^0$ | 23.87932% |
| *E. coli* | P1 [6] | 23.94 (0.43) | 22.41 (0.22) | 1.5 | 0.35 | 0.00% |
| *E. coli* | Zymo HostZero (C1) | 24.57 (0.34) | 25.34 (0.65) | -0.8 | $1.6 \times 10^0$ | 37.93000% |
| *E. coli* | Molysis Complete 5 (C2) | 23.73 (0.67) | 23.27 (0.55) | 0.5 | 0.74 | 0.00% |
| *E. coli* | P2 [7] | 22.18 (0.28) | 22.84 (0.73) | -0.7 | $1.5 \times 10^0$ | 31.14783% |
| *E. coli* | P3 [8] | 22.18 (0.28) | 21.39 (0.24) | 0.8 | 0.58 | 0.00% |
| *P. aeruginosa* | M-15 (this study) | 23.09 (0.46) | 24.26 (0.66) | -1.2 | $2.1 \times 10^0$ | 53.08692% |
| *P. aeruginosa* | P1 [6] | 21.84 (0.32) | 23.53 (0.27) | -1.7 | $3.3 \times 10^0$ | 69.37233% |
| *P. aeruginosa* | Zymo HostZero (C1) | 25.98 (0.23) | 26.70 (0.17) | -0.7 | $1.7 \times 10^0$ | 39.94829% |
| *P. aeruginosa* | Molysis Complete 5 (C2) | 25.03 (0.38) | 35.31 (1.79) | -10.3 | $6.8 \times 10^2$ | 99.85400% |
| *P. aeruginosa* | P2 [7] | 23.62 (0.46) | 24.09 (0.23) | -0.5 | $1.4 \times 10^0$ | 30.04768% |
| *P. aeruginosa* | P3 [8] | 23.56 (0.44) | 23.43 (0.44) | 0.1 | 0.92 | 0.00% |
| *S. aureus* | M-15 (this study) | 20.59 (1.15) | 22.49 (0.32) | -1.9 | $4.5 \times 10^0$ | 77.91082% |
| *S. aureus* | P1 [6] | 21.94 (1.06) | 22.74 (0.40) | -0.8 | $2.0 \times 10^0$ | 50.21738% |
| *S. aureus* | Zymo HostZero (C1) | 26.76 (0.62) | 26.07 (0.27) | 0.7 | 0.66 | 0.00% |
| *S. aureus* | Molysis Complete 5 (C2) | 22.78 (0.31) | 23.33 (0.61) | -0.6 | $1.4 \times 10^0$ | 28.46943% |
| *S. aureus* | P2 [7] | 22.78 (0.31) | 23.46 (0.23) | -0.7 | $1.6 \times 10^0$ | 38.07282% |
| *S. aureus* | P3 [8] | 22.78 (0.31) | 23.04 (0.26) | -0.3 | $1.2 \times 10^0$ | 16.75203% |

SD: Standard deviation, Ct: Cycle threshold, ΔCt: Difference in quantification cycle

### Primers, probes, and amplification parameters

To assess the depletion efficiency of the protocols, universal 16S rRNA, 18S rRNA and bacterial species-specific primers and probes were utilised [9–12]. The total volume of individual qPCR reactions was 20 µL, with 2x Rotor-Gene Multiplex PCR master mix (Qiagen, Hilden, Germany) and 2 µL DNA template. qPCR reactions were performed on a Rotor-Gene Q real-time cycler for up to 40 cycles (Qiagen, Hilden, Germany). The primer/probe sequences, concentrations and PCR cycling parameters are provided in Tab. S4.

### Benchmarking host depletion kits and protocols

A chemical host DNA depletion protocol, named M-15, was developed in this study (Fig. 1**, Supplement Protocol: M-15 mNGS**) and benchmarked alongside two commercial [Zymo-HostZERO (named C1; Zymo Research, CA, USA), MolYsis-Complete5 (C2; Molzym GmbH & Co, Bremen, Germany)] and three published host depletion protocols P1 [6], P2 [8], and P3 [7].

**Fig. 1:**
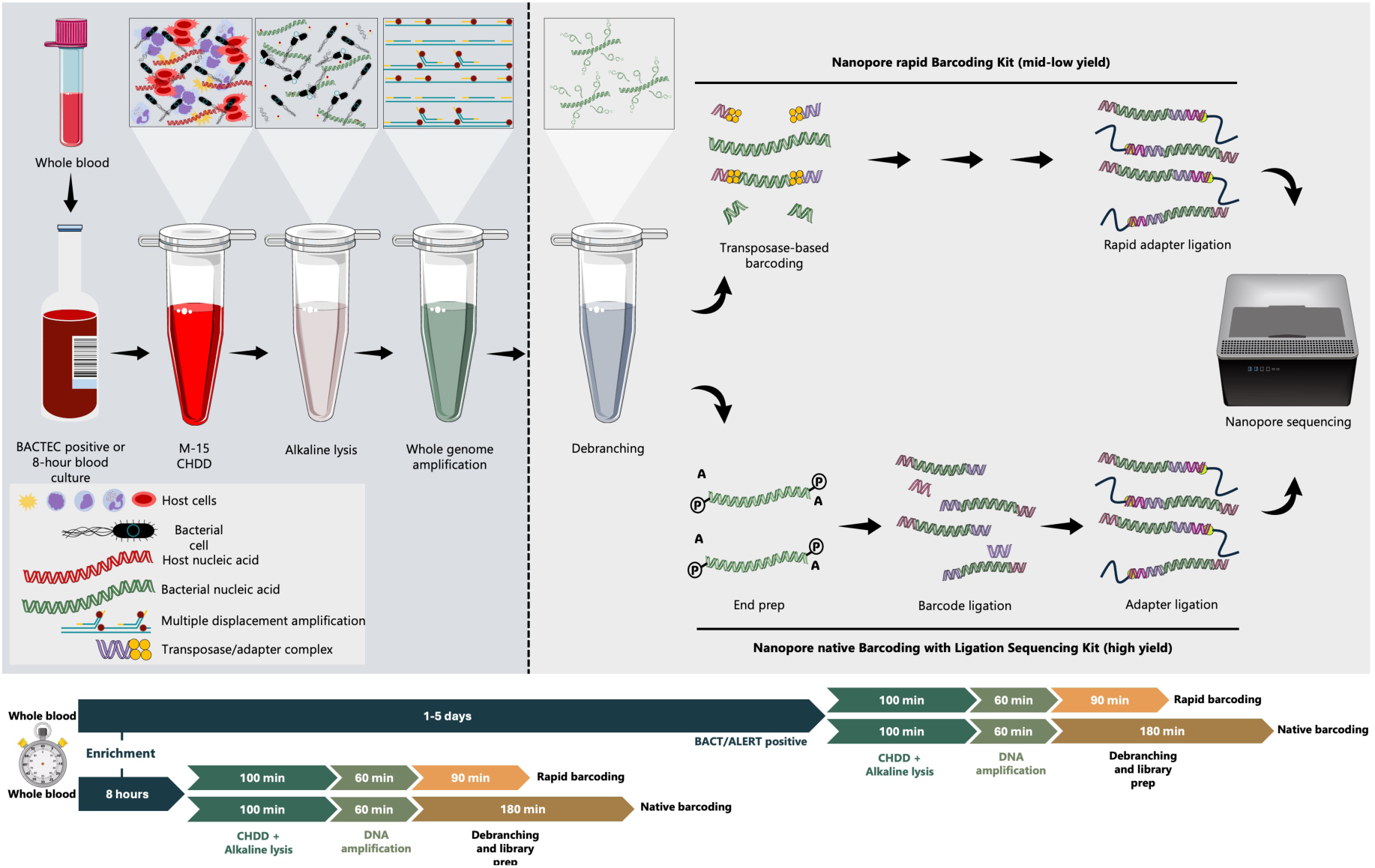
Schematic diagram depicting major steps involved in M-15 CHDD and mNGS workflow. Both BACTEC flagged positive and rapid enriched (8-hour), blood culture samples can be used as input. M-15 CHDD is the first step where host DNA is selectively depleted with saponin and DNase I treatment. Host depleted materal is then subjected to alkaline lysis and whole genome amplification with REPLI-g single cell kit. Amplified product is then debranced using T7 Endonuclease I and sequencing is performed with rapid barcoding (mid-low yield) or native barcoding (high-yield) kit.

To determine host depletion efficiency and analytical sensitivity of the workflows, serially diluted (100 to 105 CFU/mL) exponential growth phase cells of Escherichia coli (ATCC 25922), Staphylococcus aureus (ATCC 25923), and Pseudomonas aeruginosa (ATCC 27853) strains were spiked in 400 µL sterile whole blood (human and sheep) and processed to deplete host cells/DNA with all the six protocols. Following host depletion with P1, P2, P3 and M-15, DNA extraction was performed using QIAamp UCP Pathogen Mini Kit (Qiagen, Hilden, Germany) with the recommended homogenisation step by bead beating (Pathogen Lysis Tubes L, Qiagen, Hilden, Germany). For protocols C1 and C2, DNA extraction was performed using the reagents supplied with the kits. Total DNA was extracted from the same spiked blood samples without host depletion similarly as unenriched (non-CHDD) controls. Extracted DNA was tested using qPCR with the 16S rRNA, 18S rRNA, and species-specific primers/probes described above.

In addition, a subset of the simulated M-15 CHDD samples, containing 10^4^ to 10^1^ CFU/mL blood was extracted, amplified and barcoded for sequencing using ONT Rapid PCR barcoding kit (SQK-RPB004; Oxford Nanopore technologies, Oxford, UK) and sequenced on MinION R9.4.1 flow cells. Live GPU basecalling was performed in high accuracy mode (dna_r9.4.1_450bps_hac) and passed Fastq reads were analysed using Chan Zuckerberg infectious diseases (CZ ID) (v0.7; https://czid.org/) Nanopore workflow (last accessed: 07/08/2024) [13].

### Determining the contribution of host depletion on bacterial cell loss/viability

Bacterial cell loss and/or viability following host depletion was determined using *E. coli, S. aureus,* and *P. aeruginosa*ATCC strains. Approximately 30 and 3 CFU of the exponential phase cells were spiked in 400 µL sterile whole blood, and the samples were processed to deplete host DNA with three top performing protocols C1, C2, P1, and M-15. Host depleted samples were resuspended in PBS and plated on nutrient and blood agar media to estimate bacterial concentrations using the method described by Miles & Misra [14]. The same samples were plated directly without host depletion as controls. All the inoculated plates were incubated overnight at 37°C, and the number of visible colonies was recorded the next day.

### Rapid M-15 mNGS workflow tested on culture enriched specimens

To determine the efficiency of M-15 mNGS protocol on culture enriched samples, six culture enriched suspected BSI samples were tested initially. Three samples flagged positive on BACT/ALERT (bioMérieux, Marcy-l’Étoile, France) and the remaining three were reported to be culture negative. The same samples were processed directly without M-15 as no depletion controls. To minimise hands on time, rapid alkaline cell lysis was performed using 4 µL direct or host depleted samples resuspended in 1 mL PBS using REPLI-g single cell kit (Qiagen, Hilden, Germany). Multiple displacement amplification (MDA) was performed with the same kit for 60 minutes according to the manufacturer’s recommendation. Amplified MDA products were then debranched for 10 minutes at 37° C using T7 endonuclease (New England Biolabs, MA, United States) and subjected to 0.5x AMPure bead cleanup. Library was prepared using ONT Rapid barcoding kit (SQK-RBK004) with 400 ng amplified debranched DNA product/sample as an input (**Supplement Protocol: M-15 mNGS + low-medium yield protocol**). The overall sample processing time of this pilot version of the workflow is approximately 4 hours (processing 6-10 samples), including 80 minutes for host depletion, 20 minutes for DNA extraction (alkaline lysis method), 60 minutes of WGA using REPLI-g Single Cell Kit, and 90 minutes for debranching, cleanup, library preparation and flow cell loading.

To assess the efficiency of adaptive sampling (AS) with or without CHDD with M-15, rapid adapter ligated pooled libraries (six CHDD and six no CHDD) were loaded on a MinION R9.4.1 flow cell and the total number of channels (N= 512) available for sequencing were divided into two parts from the advanced option menu on the MinKNOW graphical user interface. Channels 1 to 256 were chosen for human DNA depletion using T2T-CHM13v2.0 as reference [15] and channels 257-512 for no-adaptive sampling control. High yield M-15 mNGS workflow validated with culture-positive suspected BSI specimens

An additional 27 blood culture positive samples were tested using the optimised workflow for improved sequencing yield. Chemical host depletion, DNA extraction, MDA (60 minutes) and debranching were performed similarly to the pilot protocol. Only this time, library preparation was performed using Ligation Sequencing Kit (SQK-LSK109) with native barcodes (EXP-NBD104) with 400 ng amplified debranched DNA product/sample as an input (**Supplement Protocol: M-15 mNGS + high yield protocol**). The overall sample processing time of this optimised workflow takes less than 6 hours (with 6-10 samples) including 80 minutes of CHDD, 20 minutes for DNA extraction (alkaline lysis method), 60 minutes of WGA using REPLI-g Single Cell Kit, and about 180 minutes for debranching, library preparation and loading.

To identify bacterial species, CZ ID pipeline was utilised as mentioned above with a 20% abundance cutoff (200,000 Base Per Million; BPM for CZ ID). Host-filtered contigs remaining after the 20% abundance cutoff were analysed to identify AMR determinants with ResFinder 4.5.0 [16] using the default options. In addition, host filtered fastq reads remaining after 20% abundance cutoff were mapped against bacterial reference sequences using minimap2[17] to estimate the breadth of coverage and sequencing depth. To determine the minimum sequencing yield required to identify bacterial species and AMR determinants, 100 Mbp, 50 Mbp, 40 Mbp, 30 Mbp, 20 Mbp, and 10 Mbp raw fastq reads were randomly subsampled using rasusa [18] and processed similarly with CZ ID and ResFinder.

### Rapid culture enrichment to accelerate microbiological diagnosis

A shorter culture enrichment method was tested with 16 ATCC/clinical isolates representing eight Gram negative and seven Gram positive bacterial species. Approximately 1-10 CFU of actively growing (log phase) bacterial cells were spiked in BD BACTEC™ Standard Blood Culture media (Becton, Dickinson and Company, NJ, USA) supplemented with 10 mL sterile whole blood (sheep), and then subjected to incubation. Culture bottles were taken out at 2-hour intervals over a span of 10 hours and subsequently plated on nutrient and blood agar media using the Miles & Misra method.[14] All the plates were incubated at 37°C overnight and the number of visible colonies was recorded the next day. One mL culture-enriched samples were preserved at-80°C during each sampling session to be tested later. Finally, a subset (10/16) of the 8-hour culture-enriched samples (5 Gram-positive and 5 Gram-negative species) were chosen and processed with the high yield M-15 mNGS protocol (**Supplement Protocol: M-15 mNGS + high yield protocol**). Species, host: pathogen ratio, antimicrobial resistance genes and minimum DNA yield per sample for identification were determined using the same pipelines and cutoffs mentioned above.

### Conventional tests for detecting species, AMR, and predicting AMR with sequencing

In the routine diagnostic microbiology laboratory, 5-10 mL EDTA treated blood samples collected from suspected individuals were incubated in the BACT/ALERT system for up to 5 days. Samples that flagged positive were taken out and inoculated onto appropriate agars, and organisms grown identified using MALDI-TOF mass spectrometry (Bruker Corporation, MA, USA). Phenotypic antimicrobial sensitivity was determined using disc diffusion as described by European Committee on Antimicrobial Susceptibility Testing (EUCAST) guideline [19] or automated systems, e.g. Vitek2 (bioMérieux, Marcy-l’Étoile, France).

To compare routine phenotypic sensitivity and genotypic antimicrobial susceptibility results, AST results for the BACT/ALERT positive BSI samples were compared to ResFinder 4.5.0 phenotype table results. Because ResFinder provides AMR predictions for 92 antimicrobial agents from 21 antibiotic classes, but only predicts resistance or susceptibility (not Intermediate results), only the phenotypic results common between the two (culture and sequencing) and classified as resistant or susceptible were compared. For rapid culture enriched spiked blood samples, for which AST results were not available, WGS (where available) and M-15 mNGS data of the ten ATCC/clinical isolates were compared similarly using ResFinder 4.5.0 phenotype table. Categorical agreement, major error and very major error were calculated using the following formula [20]:

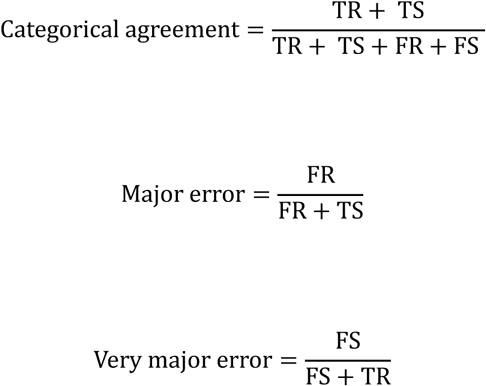

Where: TR = True resistant, TS = True susceptible, FR = False resistant, FS = False susceptible, and where phenotypic AST is taken as the gold standard.

### Statistical analysis

R studio 2023.12.1 with R base version 4.3.3 were used in combination with readxl, dplyr, reshape2, ggplot2, and gridExtra packages for performing data cleanup, formatting, analysis and generating graphs.

## Results

### Chemical depletion protocols remove unwanted host with high efficiency

Protocols M-15 and P1 removed 4.1×10^6^ (Quantification cycle ΔCt-20.52) and 5.5×10^5^ fold (ΔCt-17.45) host DNA, respectively, from the whole blood samples spiked with Escherichia coli, Staphylococcus aureus, or Pseudomonas aeruginosa ATCC strains, with depletion efficiency over 99.99%. Commercial kit C1 removed 99.82% (5.87×10^2^ fold) host, and C2, 97.93% (4.84×10^1^ fold). Approximately 81.09% and 59.19% host were removed with the protocols P2 and P3, respectively (Tab. 1). Between 5.58% (P3) and 51.63% (M-15) bacterial DNA were removed during host depletion with the six kits/protocols. Protocols with higher depletion efficiencies also exhibited increased bacterial DNA loss (Tab. 1).

Protocol C2 exhibited a strong negative bias towards *P. aeruginosa*, as >99% (ΔCt −10.27) loss was observed, possibly due to the detrimental effect of buffer CM (a selective host cell lysis buffer provided in the kit) on this bacterial species during the host lysis steps. In a recent study, where blood culture positive samples were processed with protocol C2 to deplete host and enable rapid sequencing-based species and AMR identification [20] sequencing results also indicated suboptimal accuracy of C2 mNGS for *P. aeruginosa* compared to other bacterial species, possibly indicating similar lysis bias [20].

### CHDD protocols perform poorly with samples containing low bacterial concentrations

Although the host depletion efficiency of some of the protocols, including M-15 was encouraging, we observed some bacterial loss. Therefore, we aimed to evaluate their applicability on blood samples with low bacterial concentrations (1 to 100 CFU/mL), as typically seen in BSI cases [21]. For this, we selected the top performing protocols, M-15, P1, C1 and C2, which exhibited the highest host depletion efficiency. For all three ATCC strains tested (*E. coli, S. aureus, P. aeruginosa*), qPCR analytical sensitivity of protocols M-15, P1 and C1 was 10^2^ CFU/mL. With protocol C2, qPCR was able to detect up to 10^1^ CFU/mL *E. coli* and *S. aureus*; however, like before, Ct values from *P. aeruginosa* spiked CHDD samples were consistently very high (Ct >30) across all five *P. aeruginosa* concentrations (10^0^ to 10^4^ CFU/mL) spiked in whole blood (Fig. 2A).

**Fig. 2:**
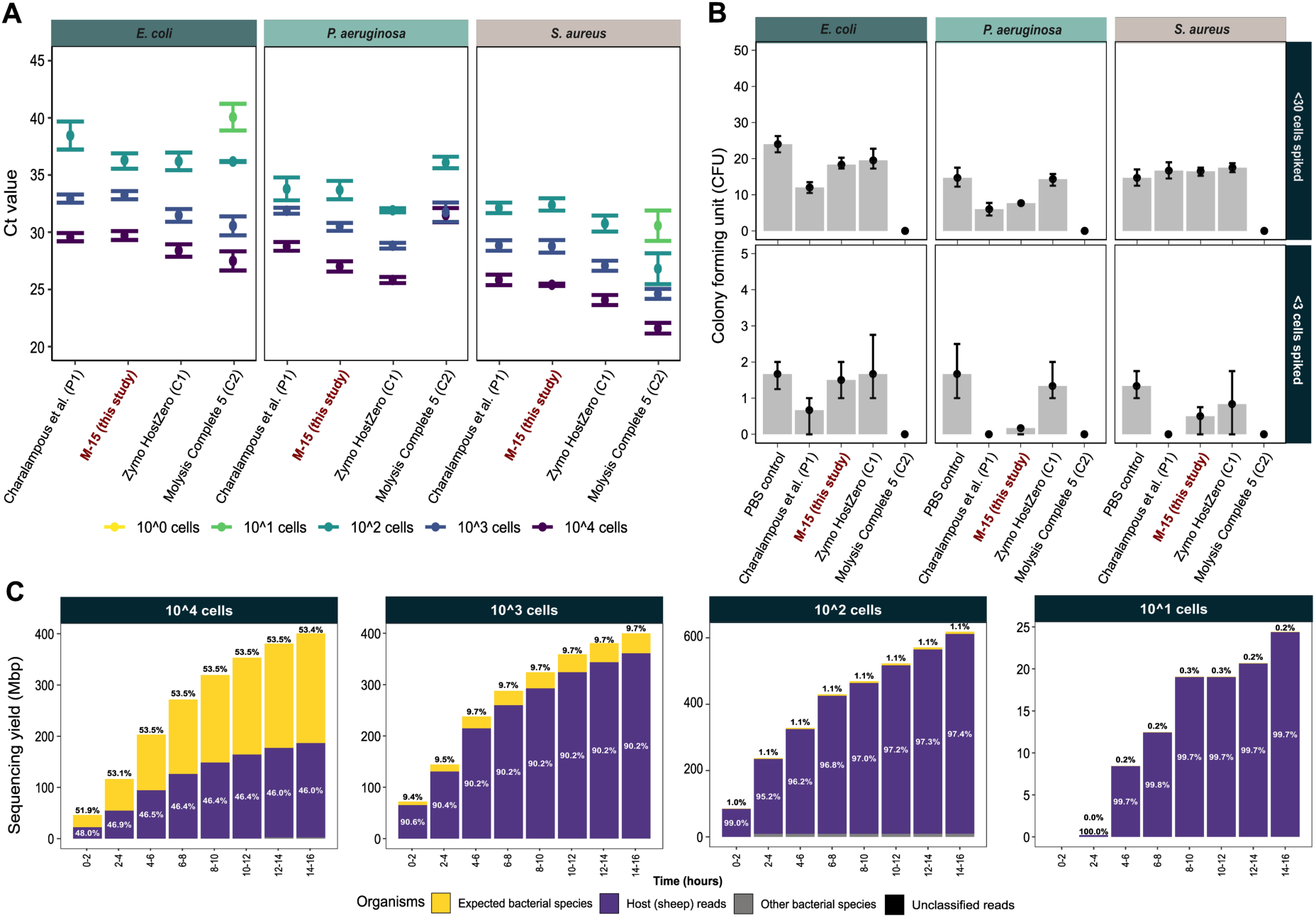
Overall performance of M-15 versus published/commercial protocols (A) qPCR Ct values (median and standard deviation) of different concentrations (10^4 to 10^0 CFU/mL – see legend) of three ATCC strains (*Escherichia coli*, *Pseudomonas aeruginosa*, and *Staphylococcus aureus*) spiked in whole blood and processed with top chemical host DNA depletion (CHDD) kits/protocols (P1, M-15, C1, and, C2) performing the best in terms of host DNA depletion. (B) Number of viable cells recovered from blood samples spiked with <30 and <3 CFU of the three ATCC strains following CHDD with P1, M-15, C1, and, C2, compared to the no CHDD/PBS control. (C) Proportion of expected bacteria initially spiked in blood for M-15 CHDD, compared to host (sheep) DNA, other bacterial species (different from the three species spiked), and unclassified reads, over 16 hours of MinION sequencing. The four facets are showing simulated/spiked bacterial concentration ranging from 10^4^ to 10^1^ CFU/mL of sheep blood.

To understand which part (chemical wash or DNA extraction) of the CHDD protocols is contributing to reduced recovery of bacterial DNA, we processed low abundant spiked samples (<30 and <3 CFU/mL) with M-15, P1, C1 and C2 protocols and confirmed viability/recovery with culture. Viable plate counts following CHDD showed protocols C1 retained 92%, M-15 76% and P1 62% of bacteria of the low initial counts of <30 cells spiked in blood (Fig. 2B). With even lower concentration (<3 cells), the overall bacterial recovery rate with protocols C1, M-15, and P1 were 72%, 43% and 13% respectively compared to the no CHDD controls (Fig. 2B). No viable bacteria were recovered using protocol C2 for any of the bacterial species or samples tested; again, possibly indicating buffer CM’s effect on their growth or viability, although *P. aeruginosa* was the only species (specifically ATCC 27853) tested in this study that displayed suboptimal performance with downstream molecular assays (Fig. 2B).

In order to estimate the proportions of expected bacterial DNA (i.e., bacterial species spiked initially) versus host and others (unclassified and other species), a subset of simulated M-15 CHDD samples (highest host depletion efficiency; 4.1×10^6^ fold), having 10^4^ to 10^1^ *E. coli* CFU/mL were sequenced. While the host: bacteria DNA ratio might differ between bacterial species due to the difference in their genome size, this would facilitate estimating the lowest bacterial concentration (CFU/mL) that would most likely provide consistent results with downstream molecular applications, especially sequencing. The proportion of expected bacterial (*E. coli* that were initially spiked) DNA sequence yield (mega base pairs; Mbp) in M-15 CHDD samples were 53.4% and 9.7% of total DNA for samples containing 10^4^ and 10^3^ CFU/mL bacteria respectively. For low abundant samples, bacteria represented only 1.1% (10^2^ CFU/mL) and 0.2% (10^1^ CFU/mL) of total DNA yield following >99.99% host removal with M-15 (Fig. 2C). Proportion of DNA sequence yield from expected bacterial species compared to the host remained unchanged throughout the sequencing run regardless of bacterial concentration spiked or DNA ratio (Fig. 2C). The suboptimal host: pathogen ratio in low abundant blood samples meant that sequencing-based assays may not yet provide satisfactory results even with CHDD, unless an additional enrichment/depletion step is included pre/post DNA extraction to further improve bacterial proportions, and a more robust and sensitive DNA extraction method is benchmarked and validated.

### M-15 mNGS and adaptive sampling significantly improve bacterial proportions

Due to suboptimal performance of M-15 mNGS observed with low bacterial concentrations directly in blood samples, three BACT/ALERT positive (BCP01-BCP03) and three negative (BCN01-BCN03) culture enriched samples from suspected BSI cases were initially tested with a rapid version of M-15 CHDD protocol (Fig. 1).

Length of the sequencing reads varied significantly among organisms, with expected bacterial species (species that were identified with culture-based methods) having the highest median read length (1,957 bp); this was 8.15x longer than the unclassified reads (240 bp), 3.52x longer than human reads (552 bp) and 1.7x longer than other bacterial reads (mixed bacterial species not expected from the sample; 1,145 bp) (Fig. 3A). The read length difference between host and bacteria could be the result of CHDD, as host DNA was digested with DNase I following selective lysis.

**Fig. 3:**
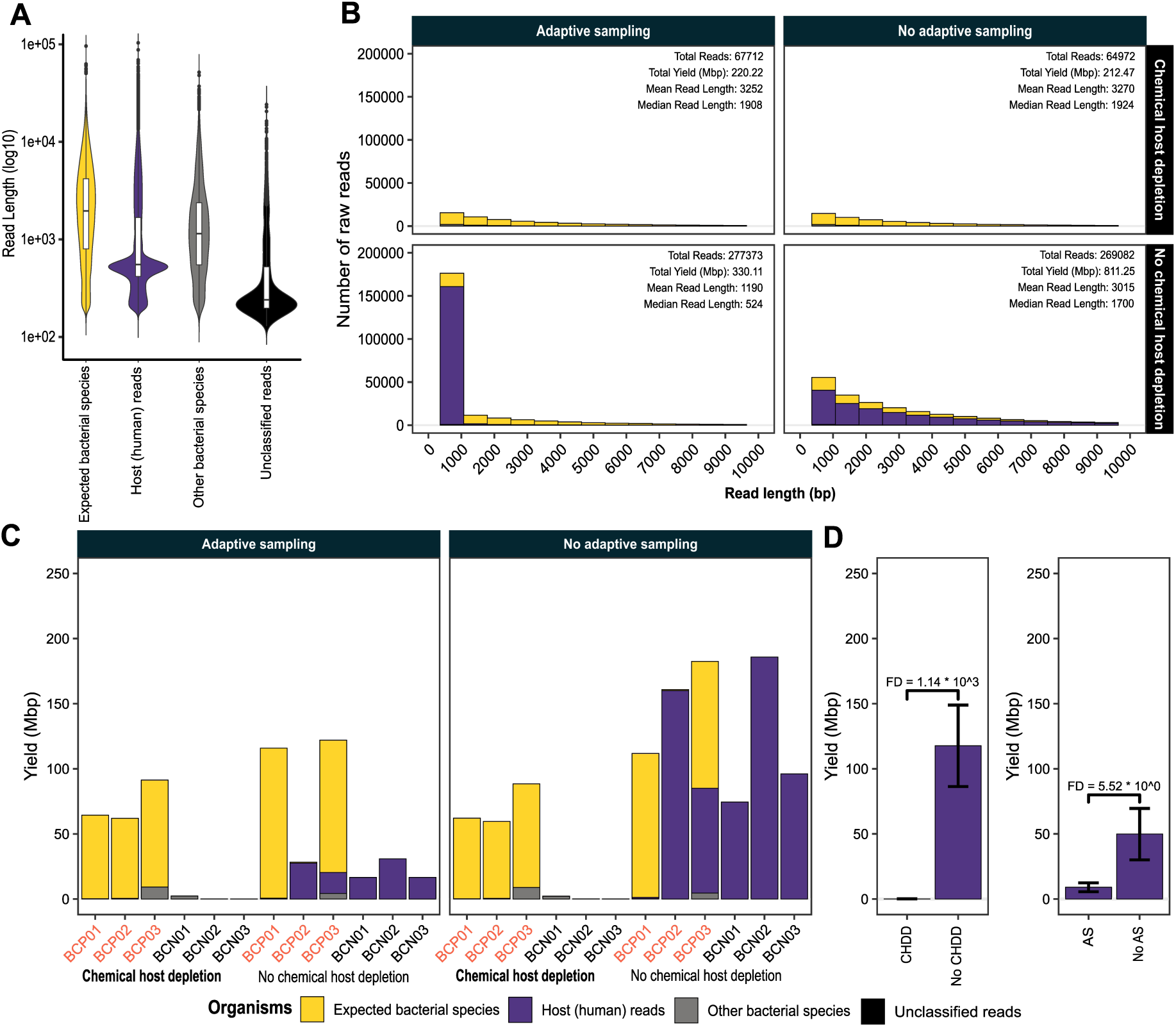
Host depletion efficiency of M-15 mNGS and adaptive sampling (AS), versus no chemical host DNA depletion (CHDD) and no AS, tested with patient samples (A) Violin plot showing size distribution of sequencing reads aligned to expected bacterial species, host (human), other bacterial species, and reads for which there were no matches within the blast Nr/Nt database. (B) Histogram highlighting the number of raw sequencings reads from expected bacteria, host, others (other bacteria or unclassified). Coloured facets subdividing the raw reads further to distinguish read length variation by AS, no AS and CHDD and no CHDD. (C) Overall sequencing yield of CHDD and no-CHDD samples sequenced, comparing AS and no-AS. Stacked coloured bars highlight the proportion of DNA aligned to host, bacteria and others/unclassified species (see legend). Blood culture positive (BCP) samples are shown in red text, while blood culture negative (BCN) samples are shown in black. (D) Depletion efficiency (fold difference, FD) in DNA yield (Mbp) of sequences from host with CHDD and AS compared to no-CHDD and no-AS.

Overall yield in the adaptive sampling (AS) channels (1-256) was 1.86x lower (550.33 Mbp AS vs 1023.72 Mbp no-AS) than no-AS (channels 257-512), although the total number of reads for AS and no-AS samples was almost the same, with 345,085 and 334,054 reads, respectively. Median read length for AS channels was 3.2x shorter than the no-AS, and host constituted >90% of the reads that were smaller than 1kb in AS channels (Fig. 3B). This illustrates that AS influenced the length of sequenced fragments as a result of its limited ability to deplete shorter host reads (≤1 Kbp), which were highly abundant in the directly extracted no-CHDD samples.

Samples processed with M-15 CHDD had 1.14 ×10^3^ times less human DNA compared to the no CHDD controls (based on sequencing yield in Mbp; Fig. 3D). Samples sequenced using AS had 5.52-fold less human DNA compared to the controls; however, no bacterial enrichment was observed (Fig. 3C, 3D). This highlights the importance of host depletion even in positively flagged culture-enriched specimens, as it may lead to lower sequencing cost per sample, and more importantly, reduce the sequencing time required to reach a desired yield for determining the bacterial species responsible for the BSI.

### M-15 mNGS accurately predicts mono-bacterial species in BACT/ALERT positive samples

Because suboptimal sequencing yield was observed when debranched M-15 CHDD Multiple Displacement Amplification (MDA) samples were initially processed (rapid M-15) using the rapid barcoding kit (Fig. 1), it was decided to utilise the Ligation sequencing kit with native barcodes (Fig. 1). Additional 27 culture (BACT/ALERT) positive BSI blood samples were processed with the high-yield M-15 mNGS protocol.

Overall, blood-culture positive (BCP) samples subjected to CHDD had 152.4x higher DNA concentration compared to culture negative (BCN) samples (BCP CHDD 509.02 ng/µL versus BCN CHDD 3.34 ng/µL, as measured using Qubit) after 60 minutes MDA (Fig. 4A, Tab. S1). However, directly extracted and amplified blood culture negative samples yielded 753 ng/µL DNA on average following 1-hour MDA with REPLI-g kit (Tab. S1). Four bacterial species with high GC content, *Brevibacterium luteocum* (GC 67.8%)*, Micrococcus luteus* (GC 74.0%)*, Stenotrophomonas maltophilia* (GC 66.7%) and *Achromobacter xylosoxidans* (GC 64.0%) yielded low DNA (15.7x less than other species), with an average of 32.23 ng/µL after 1-hour MDA; however, this was still 9.64x higher than BCN samples following CHDD (Tab. S1). The potential reason behind low yield in species with high GC%could involve stable secondary structure or denaturation difficulties as highlighted in some studies [22, 23].

**Fig. 4:**
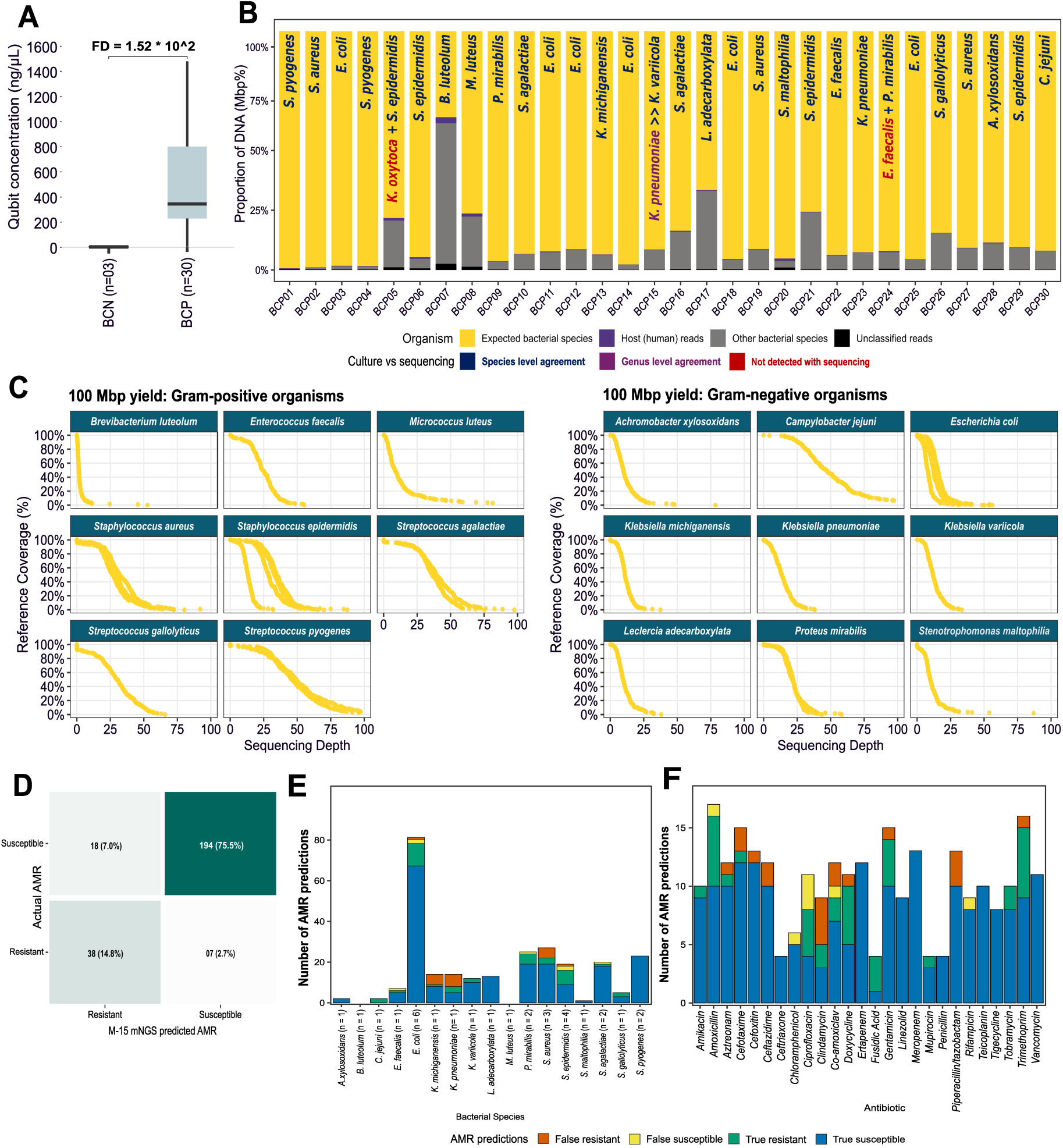
Species and antimicrobial resistance prediction accuracy of M-15 mNGS (A) DNA concentration (ng/μL) of three blood culture negative (BCN01-BCN03) and 30 blood culture flagged positive (BCP) samples, following M-15 chemical host DNA depletion (CHDD), and whole genome amplification with the REPLI-g kit. Quantification was performed with Qubit Broad Range dsDNA assay. (B) Proportion of expected bacterial DNA (yellow bar chart; confirmed by culture) versus all other reads following M-15 CHDD and MinION sequencing with native barcoding kit. BACT/ALERT positive samples that have species-level agreement with culture have text labelled dark blue, those with genus-level agreement are purple, and those not detected by sequencing are labled red. (C) Cumulative depth for eight Gram-positive and nine Gram-negative organisms from 30 BACT/ALERT positive blood culture samples following M-15 mNGS, with 100 Mbp sequencing yield per sample. (D) Number and proportion of true positive, true negative, false positive and false negative AMR predictions with M-15 mNGS compared to the standard culture-based AST prediction for 30 BACT/ALERT positive blood culture samples. (E) Number and proportion of ResFinder true positive, true negative, false positive and false negative AMR predictions with M-15 mNGS by bacterial species from the 30 BACT/ALERT positive blood culture samples tested in this study. (F) Number and proportion of true/false AMR predictions with mNGS for the same culture-positive samples categorised by antibiotics tested. Prediction of AMR profile with mNGS was done only using the resistance and susceptibility results from antibiotics that were present in both phenotypic and genotypic datasets (n=257).

With CZ ID, using a cutoff of 20% abundance (200,000 Bases Per Million: BPM) successfully removed all bacterial species identified except those detected using culture-based methods. Using this cutoff, sequencing correctly identified 28/28 mono-bacterial species, 3/3 negative samples and 2/4 multi-bacterial species found in two samples (Fig. 4B, S1 A).

Initially, it was assumed that the two species (*K. oxytoca* and *E. faecalis*) in two mixed samples (BCP05, and BCP24) were missed due to bacterial cell lysis during CHDD (Fig. 4B). However, some of the mono-bacterial samples tested in this study contained the same bacterial genera/species and no negative bias was observed with sequencing following CHDD. Also, all the CHDD flagged positive samples (n=30, species=17) were cultured overnight to confirm viability, and only one (*M. luteus*) showed visible growth impairment on agar plates (Tab. S1). Therefore, it is likely that these two organisms were missed with sequencing due to their relatively low abundance compared to the other species present in the sample. For one sample (BCP15), *Klebsiella pneumoniae* was identified with culture but *Klebsiella variicola* with mNGS; however, the species was later confirmed to be *K. variicola* with 16S rRNA amplicon sequencing. Overall, expected bacterial species constituted 88.7% of total sequencing yield versus human which represented only 0.46% total DNA (Fig. 4B).

To understand the breadth of coverage across the bacterial genomes with 100 Mbp sequencing yield per CHDD sample, 100 Mbp fastq reads subsampled with rasusa were analysed. For every 100 Mbp DNA sequenced per sample, 96.76% and 99.48% coverage could be achieved with at least 1x depth for the Gram-positive and Gram-negative organisms tested, respectively. For 5x and 10x depth, Gram positive organisms had a mean genome coverage of 87.89% and 78.29% and Gram-negatives 95.28% and 78.38%, respectively (Fig. 4C). For all the 30-blood culture positive samples tested (BCP01 to BCP30), >20% abundance cutoff accurately identified the present bacterial species with as low as 10 Mbp sequencing yield per sample (Fig. S1 A).

### Rapid culture enrichment can facilitate same-day microbiological identification

When a blood culture is flagged as positive, it contains approximately 10^7^ to 10^9^ bacterial cells/mL of culture broth[24], which is more than the input concentration (≥10^4^ CFU/mL) required for the M-15 mNGS to work reliably, as observed during our initial experiments (Fig. 2C). This meant that a shorter culture enrichment step could further accelerate the microbiological diagnosis, and M-15 mNGS could be utilised even before a sample flags positive. However, given the variety of bacterial species involved in BSI and differences in their growth rate, we aimed to understand the minimum incubation period required to reach that threshold with a representative range of clinically important bacterial species (n=15).

Four of 15 bacterial species tested (*Streptococcus agalactiae, Acinetobacter baumanii, Klebsiella pneumoniae* and *Escherichia coli*) reached 10^4^ CFU/mL, the optimal concentration for M-15 mNGS, under 6 hours of culture enrichment (Fig. 5A). With 8-hour incubation, 12/15 species reached the desired threshold of 10^4^ CFU/mL. The remaining three species, *Staphylococcus aureus, Streptococcus pneumoniae* and *Pseudomonas aeruginosa*, were slightly lagging, reaching approximately 6.04×10^3^ CFU/mL on average with 8-hour enrichment. All the species (15/15) tested reached/crossed the threshold under 10 hours of culture enrichment (Fig. 5A).

**Fig. 5:**
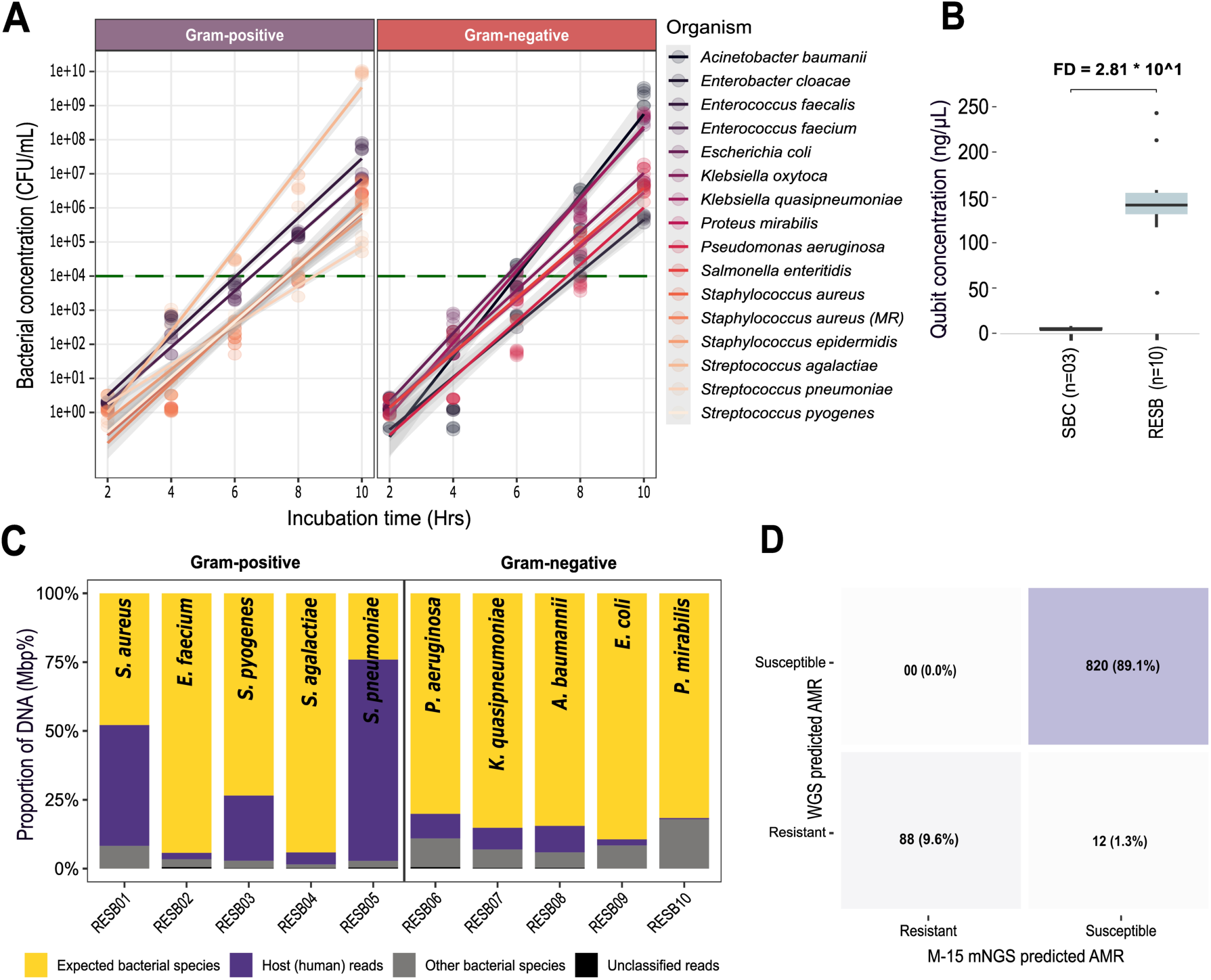
Rapid culture enrichment: species covered, overall accuracy in species and antimicrobial resistance prediction. (A) Time to reach 10^4 colony forming units (CFU)/mL (green line; optimal concentration for chemical host DNA depletion (CHDD) with culture-based bacterial enrichment prior to M-15 CHDD and mNGS, tested with the top 15 bacterial species causing bloodstream infection. (B) DNA concentration (ng/μL) of the three strerile culture-enriched blood (SBC01-SBC03) and ten rapid (8-hours) culture enriched spiked blood samples, following M-15 CHDD, and whole genome amplification with the REPLI-g kit. Quantification was performed with Qubit broad range assay. (C) Proportion of expected bacteria (yellow; spiked initially) versus all other reads following M-15 CHDD and MinION sequencing. Species, genus and non-agreements from rapid culture-enriched samples are highlighted with the same colored labels as before. (D) Number and proportion of ResFinder true positive, true negative, false positive and false negative AMR predictions with M-15 mNGS compared to the whole genome sequencing (WGS) results of the same isolates/strains. MR: methicillin-resistant *In silico* prediction of AMR is promising but requires improvement for clinical use The accuracy of AMR prediction for the BACT/ALERT positive samples was determined comparing the ResFinder phenotype antibiotic data with the phenotypic culture-based antimicrobial susceptibility testing (AST) results (n= 257). For all 30 culture positive samples, mNGS exhibited 90.32% (232/257) categorical agreement overall, with 8.49% (18/212) major error and 15.55% (7/45) very major error (Fig. 4D). Among the 17 bacterial species, discrepancies in resistance calls for *K. michiganensis* (5/18), *K. pneumoniae* (6/18), and *S. aureus* (5/18) contributed to the highest number of major errors, and *E. coli* (2/07) and *S. epidermidis* (2/07) towards very major error (Fig. 4E). Of 27 antibiotics for which results were compared, very major error for ciprofloxacin was most common (3/7), possibly due to the relatively low accuracy of ONT reads and the frequent association of resistance to fluoroquinolones with mutations in quinolone resistance determining region (QRDR) of DNA gyrase and topoisomerase encoding genes, as opposed to resistance encoding gene presence/absence in other antibiotics.[25] Clindamycin (4/18) and piperacillin/tazobactam (3/18) were the top two antibiotics exhibiting major errors (Fig. 4F).

To identify proportions of expected bacteria obtained using M-15 mNGS after 8-hour rapid culture enrichment, three sterile blood culture samples and a subset of 10 bacterial species (5 Gram positive and 5 Gram negative, including the three slow-growing species tested earlier) were selected and processed with this approach (Fig. 5A, C). One potential costly drawback with the rapid enrichment approach could be the necessity to blindly sequence all blood cultures if the positive and negative samples could not be differentiated prior to sequencing. However, following CHDD and 60 minutes of MDA, the rapid culture-enriched spiked blood samples (RESB, n=10) showed 28.16x higher dsDNA concentration (154.6 versus 5.49 ng/µL) on average compared to sterile human blood enriched as negative controls (SBC, n=3), providing a unique signature for the detection of positive samples prior to sequencing (Fig. 5B, Tab. S2).

Following sequencing, expected bacteria represented 89.4% of total DNA yield on average for 7 samples that reached 10^4^ CFU/mL under 8-hours versus 50.06% for the remaining 3 samples having relatively slow-growing species. All samples combined, 75.4% of the total DNA yield matched with the bacterial species initially used for spiking and enrichment and 17.64% with host species (Fig. 5C). Similar to BACT/ALERT positive samples, >20% abundance threshold successfully identified 10/10 bacterial species sequenced to validate the rapid enrichment protocol (Fig. 5C). Also, like before, as low as 10 Mbp yield (subsampled) was sufficient to accurately identify bacterial species with the abovementioned cutoff (Fig. S1 B).

Due to the smaller number of ATCC strains and clinical isolates (n =10) tested, and unavailability of phenotypic AST data for a few clinical isolates, AMR predictions of M-15 mNGS with the rapid culture method were compared to whole genome sequencing (WGS) of pure bacterial strains predicted AMR of the same strains/species using ResFinder (n = 920 predictions). Overall categorical agreement for M-15 mNGS versus WGS was 98.7% (908/920) and mNGS exhibited 89.1% true susceptible, 9.6% true resistant, 1.3% false susceptible and no false resistant predictions (Fig. 5D). *P. aeruginosa* was the only species for which sequencing results did not match perfectly with its reference genome (12/92 predictions) and therefore was the only source of false susceptibility, possibly due to its high GC content leading to inefficient/uneven DNA amplification with the REPLI-g kit (Tab. S2).

For both BACT/ALERT positive blood culture and rapid enriched spiked blood M-15 CHDD blood, ≥50 Mbp sequencing yield was sufficient to predict AST profiles, with the accuracy almost the same as having a sequencing yield of >100 Mbp per sample (Fig. S2, S3).

## Discussion

Timely identification of pathogens and their antimicrobial resistance profile is crucial in BSI because more targeted and effective antibiotic therapy can minimise the progression and severity of the disease and therefore lead to improved patient outcomes. In addition, reduced prescription of broad-spectrum antibiotics may help preserve their effectiveness, limit the emergence of new antibiotic-resistant strains and protect the valuable microbiota that plays an important role in maintaining overall nutrition and health [26]. Rapid sequencing-based methods, especially in combination with selective chemical depletion approaches, have the potential to reduce turnaround time and therefore lessen detrimental outcomes. In working towards this goal, this study initially assessed the effectiveness of five published and commercial, one in-house and one computation-based host depletion strategies.

As a first step, we compared the effectiveness of an in-house host depletion method (M-15) with five published and commercial approaches, as well as a computation-based host depletion strategy. The host depletion efficiency of most of the CHDD workflows was outstanding (particularly for M-15 and P1). However, we also noticed a correlation between depletion efficiency and loss of bacterial DNA with some protocols (e.g., C2) showing significant negative bias towards some species. For samples with low bacterial abundance, the proportion of bacterial DNA was insignificant, ≤1% of total DNA, even after M-15 CHDD, which can remove >99.99% host DNA. However, bacteria represented >50% of total DNA sequence yield for the CHDD samples containing ≥10^4^ CFU/mL bacteria. This meant additional enrichment/depletion was necessary to get improved proportions of the sequencing yield that is bacterial. As a simple and cost-effective alternative, we realised that a culture-based enrichment step prior to CHDD and sequencing might improve DNA extraction efficiency/robustness and proportion of bacteria in the sample simultaneously.

Therefore, we initially tested BACT/ALERT positive samples with two versions of M-15 mNGS protocols. As confirmed during the initial phase, the proportion of host: bacterial DNA sequenced was improved significantly with CHDD and AS. However, the sequencing yield was suboptimal when CHDD whole genome amplification (WGA) was paired directly with the Rapid Barcoding Kit (not systematically tested). The best output was observed when debranched MDA products were barcoded using the Ligation Sequencing Kit, although it requires additional hands-on time and potentially pricier reagents for library preparation.

Both versions of the workflows have their own strengths and limitations; for example, the rapid workflow requires less hands-on time but may result in relatively low sequencing yield, leading to higher sequencing cost per sample (Fig. 1). In contrast, the high-yield workflow would reduce sequencing cost by providing greater output with only 90 additional minutes for sample processing (Fig. 1). Several studies have already highlighted this difference;[27, 28] consequently, users may choose the workflow that best suits their need/budget.

Although there are multiple bioinformatic pipelines available for mNGS-based species identification, only a few have been developed/standardised for diagnosing bloodstream infection. The aim of this study was therefore not to benchmark these pipelines; instead, we assessed the utility of one of the well-regarded workflows, CZ ID, in combination with M-15 mNGS. With the 20% (200,000 BPM) abundance cutoff, sequencing accurately identified 3/3 negative, 28/28 mono-bacterial and 2/4 multi-bacterial species reported in two samples, with >88% DNA classified as the expected species. To determine minimum sequencing yield required to accurately identify bacterial species, we randomly subsampled the reads under the established assumption (Fig. 2C) that the proportion of host: bacteria remains consistent throughout a sequencing run [29]. The results demonstrated that the species prediction accuracy with the given cutoff remains nearly identical even with 10 Mbp yield per sample. Therefore, it is not entirely necessary to generate hundreds of Mbp reads/sample to cover ∼100% of the genome with >20x depth for patient management unless the aim is to utilise this data for hospital outbreak investigation.

With the rapid culture enrichment, only a few species tested reached/crossed ≥10^4^ CFU/mL threshold under 6 hours, and at least 8 hours of incubation was required to get more comprehensive representation of critically important Gram-positive (n=7) and Gram-negative (n=8) bacterial pathogens. M-15 CHDD and sequencing of a subset of rapid (8-hour) enriched samples (5 Gram-positive and 5 Gram-negative) further confirmed our hypothesis: sequence reads from expected bacteria represented >75% of total yield, and similar to BACT/ALERT positive samples, only 10 Mbp reads accurately identified species with the previously assigned abundance threshold.

Regarding AMR prediction, it is worth highlighting that not all expected molecular mechanisms of antibiotic resistance have been determined [30]. Additionally, a mere presence or absence of genes and/or mutations does not always correlate to phenotypic resistance or susceptibility [30]. Therefore, at present, it is less likely that a sequencing-based approach would match culture-based phenotypic results with 100% accuracy for all the bacterial species commonly identified in BSI. Like species identification, several workflows can predict AMR using mNGS data. The underlying principles, databases and/or accuracy can vary among these workflows for AMR identification [31]. Therefore, it was decided to test ResFinder, a well-regarded workflow, with M-15 mNGS, as benchmarking several workflows/bioinformatic pipelines was not within the scope of this study. M-15 mNGS combined with ResFinder exhibited overall 90.3% accuracy 8.49% major and 15.55% very major error with BACT/ALERT positive samples. These proportions closely match with some of the other studies that utilised culture enriched samples for metagenomic sequencing, although very major error rates with M-15 mNGS were found to be marginally lower [20].

With rapid culture-enriched samples, WGS data was used as comparator. Although this is less ideal than comparing with culture-based phenotypic results, we wanted to understand how closely mNGS results matched those of reference sequences of bacteria that were used for spiking. With 920 ResFinder AST predictions, mNGS showed 98.7% categorical agreement, and only 1.3% false susceptibility, primarily due to *P. aeruginosa*, the only species exhibiting discordant results. Similar suboptimal results with *P. aeruginosa* were also observed in the aforementioned study [20]. However, in our case, this likely stemmed from inefficient/uneven DNA amplification due to high GC content of *P. aeruginosa* (66.6%), as we did not observe any lysis/viability issues with culture when CHDD samples were plated on agars (Tab. S2).

Considering the overall performance, it seems more work is required in terms of wet/dry lab to make AMR predictions more accurate (e.g., Q30 reads and/or utilizing machine learning models [32]) to be accepted as a replacement of blood culture and phenotypic assays.

However, as an addition to existing methods, M-15 mNGS at its current stage still holds promise to inform treatment in comparison to empirical antibiotic therapy. For clinical management of BSI or infection in general, the accuracy of empirical treatment can range from 20% to 80% [33–35]. This means that a high proportion of cases receive inappropriate antibiotics until more targeted therapy is administered utilizing the phenotypic AST results (conventionally available in 48-72 hours), potentially risking detrimental outcomes for critically ill patients [34]. With the rapid availability of bacterial species and AMR predictions that surpass empiric prescription and therapy, mNGS can significantly refine the choice of antimicrobials, increasing the likelihood of favourable outcomes. That being said, thorough clinical validation would be required for rapid M-15 mNGS to be recommended for clinical implementation.

A possible costly drawback when implementing the rapid enrichment approach was the necessity to blindly sequence all blood cultures if the culture positive and negative samples could not be differentiated prior to sequencing. However, when compared to the negative blood samples, both BACT/ALERT positive and rapid enriched spiked M-15 CHDD blood had 152x and 28x higher dsDNA concentration following MDA, meaning any negative samples could potentially be filtered out prior to sequencing if their Qubit-measured DNA concentration provides a differentiable signature, as we observed following CHDD and WGA (Tab. S1, S2). However, the exact cutoff must be determined site-wise (different clinical settings or in different laboratories) with more negative patient samples before implementation in a clinical setting. It is important to note that this differentiation is only possible with rapid CHDD as direct extraction and amplification would not provide a discernible characteristic for negative samples due to the overwhelming abundance of host DNA masking the unique signature from the bacteria (Tab. S2). This approach may incorrectly flag some slow growing and/or bacterial species with high GC% as negative. However, because M-15 mNGS is currently intended to complement existing technologies rather than replacing them, species that might be missed would still be captured with traditional culture-based methods.

It is important to recognise that this study had several limitations. Since we only had access to different volumes of surplus EDTA treated blood samples from the suspected BSI cases, stored in a refrigerator for up to five days until reporting was complete, it was decided not to test the rapid enrichment workflow on patient samples. Instead, 15 clinically important bacterial species were chosen for initial validation. This is because the different volume and extended storage of the samples might alter the host/pathogen ratio, affect viability, and/or change the actual growth of the bacteria and therefore might not be an appropriate representative of freshly drawn samples.

Also, the rapid and high yield mNGS workflows were benchmarked, and a few batches of samples were initially processed using R9.4.1 flow cells, before more accurate R10.4.1 kits became widely available on the market. Therefore, it was decided to continue testing all the samples with library kits compatible with R9.4.1 flow cells [27]. We believe that the performance of M-15 CHDD would be further enhanced with the use of newer versions of kits/flow cells, as a higher proportion of ≥Q20 reads would likely allow mutations associated with AMR to be more reliably predicted.

## Conclusions

In summary, in this study, we demonstrated that M-15 CHDD paired with a rapid barcoding or ligation sequencing workflow is highly effective at species and AMR gene detection in BACT/ALERT positive and rapid culture enriched samples. This approach can deplete significant amount of host DNA and facilitate rapid bacterial/AMR identification, with a hands-on time of less than 4 (rapid barcoding kit) and 6 hours (ligation sequencing kit), plus time required to generate 10 Mbp (species)/ ≥50 Mbp (AMR) sequencing yield per sample. The time to achieve this yield can vary depending on the capacity of the flow cells used and/or the number of samples multiplexed in a sequencing run, therefore, it is up to the user to choose the appropriate flow cell and/or number of samples based on their needs. However, we demonstrated that with 8-hour culture enrichment, M-15 mNGS can predict causative bacterial species and accurately detect AMR determinants within 13-15 hours of sample collection, and thus has the potential to improve patient outcomes in BSI.

## Abbreviations

AMR: Antimicrobial resistance

AS: Adaptive sampling

AST: Antimicrobial susceptibility testing

ATCC: American Type Culture Collection

BSI: Bloodstream infection

BPM: Bases per million

CFU: Colony-forming units

CHDD: Chemical host DNA depletion ds

DNA: Double-stranded DNA

EUCAST: European Committee on Antimicrobial Susceptibility Testing

FDA: Food and Drug Administration (USA)

GC%: Guanine-cytosine content percentage

mNGS: Metagenomic next-generation sequencing

Mbp: Megabase pairs

MDA: Multiple displacement amplification

NHS: National Health Service (UK)

ONT: Oxford Nanopore Technologies

PBS: Phosphate-buffered saline

qPCR: Quantitative polymerase chain reaction

RBC: Red blood cells

WGA: Whole-genome amplification

WGS: Whole-genome sequencing

## Ethical statement

This study was reviewed and approved by the University of Glasgow College of Medical, Veterinary & Life Sciences Ethics Committee (Project No: 200210015) and UK National Health Services (NHS) Greater Glasgow and Clyde (R&I reference: GN19ID331). No personal information from the patients and healthy volunteers was collected and used in this study. Human DNA reads obtained from mNGS were excluded or not utilised for any bioinformatics analyses beyond quantifying the number of hosts reads generated.

## Availability of data and materials

All the raw sequencing reads (fastq.gz: host excluded) analysed and presented in this study are available on European Nucleotide Archive under the study accession “PRJEB80726”.

## Authors’ contributions

Conceptualisation of this study was carried out by TF and MSIS. Data generation, cleaning, management, formal analysis, and initial draft was written by MSIS. Project investigation, experimental method selection, clinical sample collection, formal review and coordination was done by TF, KB, KO, PE, MEM, CW, MF, and MSIS. TF (RS) and MSIS (LKAS) acquired funding for this project.

## Data Availability

All data produced in the present study are available upon reasonable request to the authors

## Acknowledgements

We would like to thank Kareen Macleod for generously providing the reference isolates used in this study. We also want to express our gratitude to Ryan Carter and Ângelo Mendes for their help and input on this project.

## Supplementary Figures, tables and protocols

**Fig. S1:**
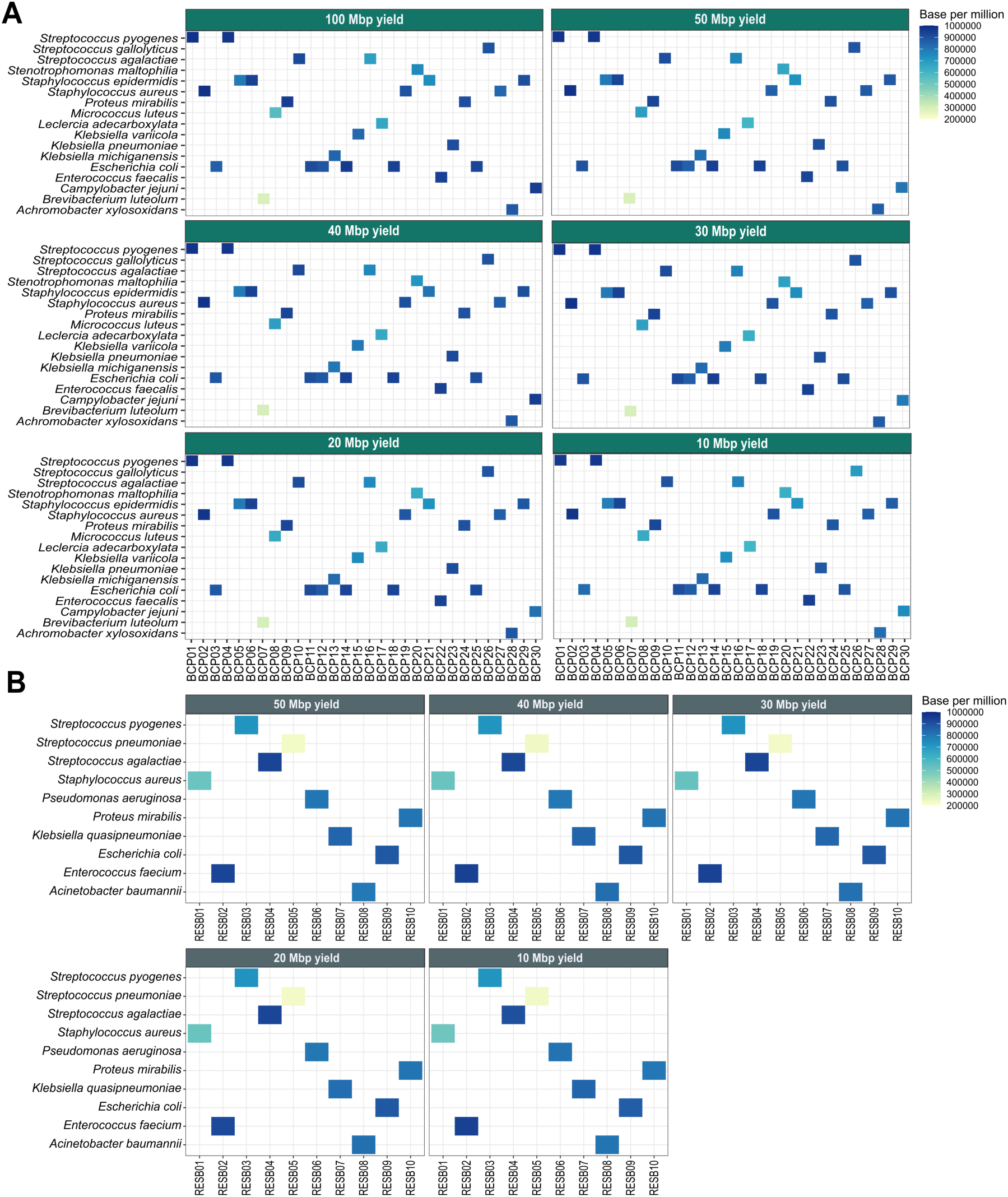
Accuracy of species prediction with varying sequencing yield (100 Mbp to 10 Mbp), identified by subsampling the (A) reads from 17 bacterial species from the 30 BACT/ALERT positive blood culture samples and (B) 10 rapid (8-hours) culture enriched spiked blood samples. Analysis was performed similarly using CZ ID with the same abundance threshold for all the species tested. Colored gradient highlights the species that passed the initial threshold filter and their abundance (base per million) in the CHDD samples that were sequenced.

**Fig. S2:**
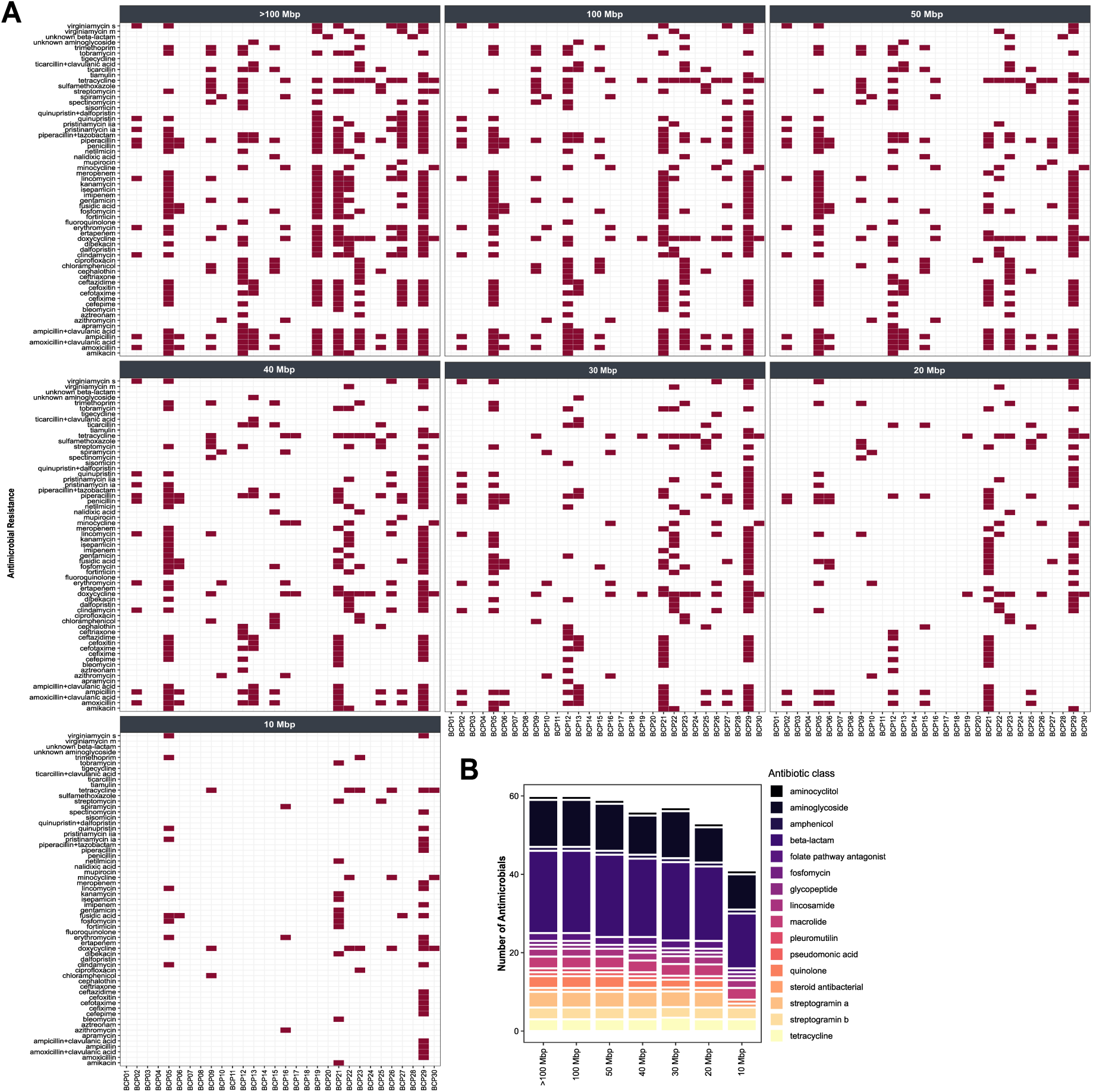
Antibiotic susceptibility profiles predicted with varying sequencing yield (>100 Mbp to 10 Mbp/sample) using subsampled reads from the 30 BACT/ALERT positive blood culture samples. (A) The heatmap is showing antimicrobial resistance profiles and (B) the barplot highlights the number of antimicrobials from different antibiotic classes identified with different sequencing yield for BACT/ALERT positive samples.

**Fig. S3:**
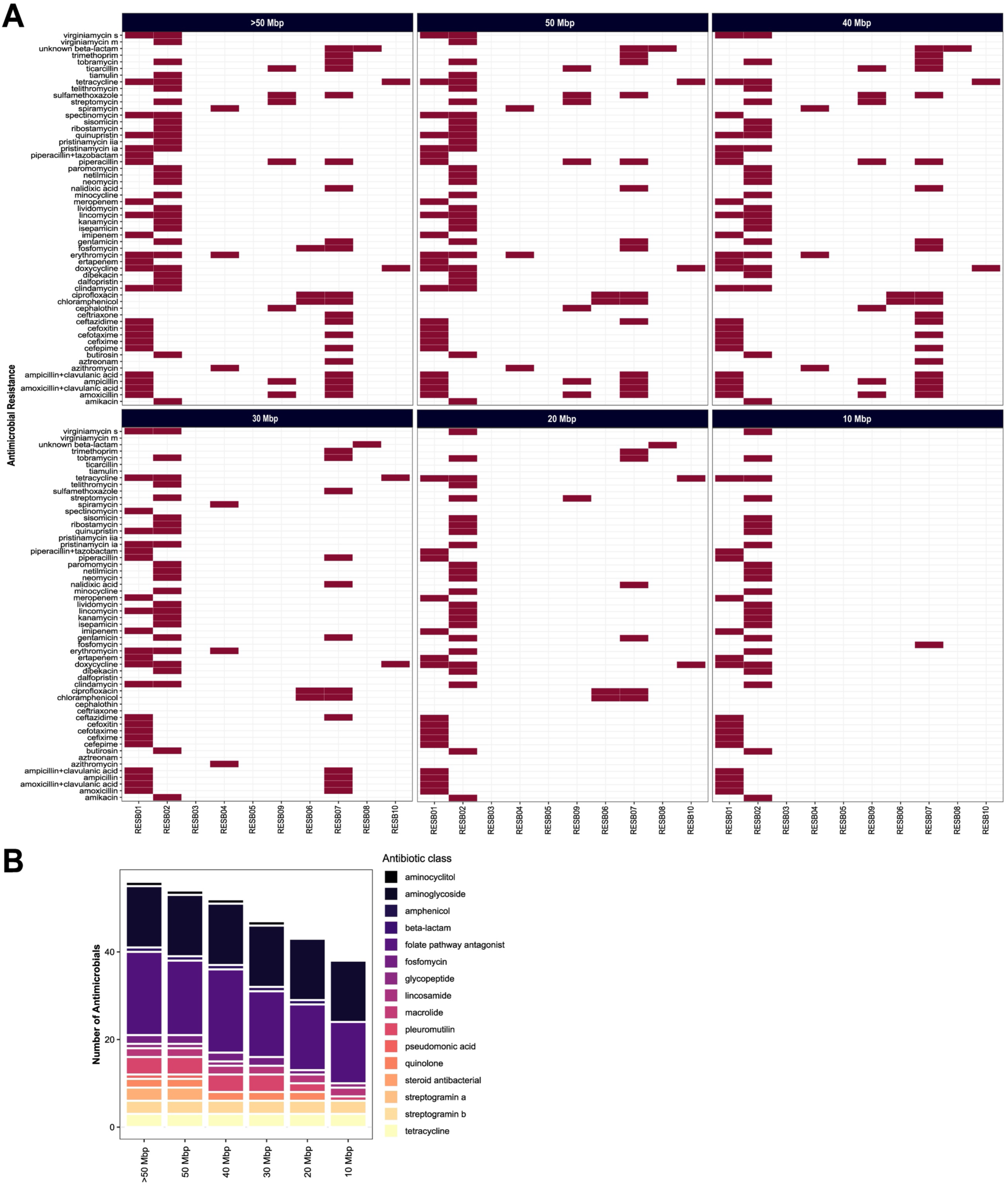
Antibiotic susceptibility profiles predicted with varying sequencing yield (>50 Mbp to 10 Mbp/sample) using the subsampled reads from 10 rapid culture enriched (8-hours) spiked blood samples. (A) The heatmap shows antimicrobial resistance profiles and (B) the barplot highlights the number of antimicrobials from different antibiotic classes identified with different sequencing yield for rapid culture enriched spiked blood samples.

**Tab. S1:** Double stranded DNA concentration of three BACT/ALERT negative (BCN01 to BCN03) and 30 BACT/ALERT positive blood culture (BCP) samples quntified by Qubit following M-15 CHDD and 60 minutes of multiple displacement amplification with REPLI-g kit. As controls, six samples (BCN01 to BCN03 and BCP01 to BCP03) were also extracted directly and amplified similarly for 60 minutes without CHDD. Viability of bacterial following M-15 CHDD was confirmed by plating the supernatant on nutrient and blood agar media.

| Sample | Accession | Organism | Culture | GC content (%) | Viability after M-15 CHDD | CHDD samples DNA (ng/μL) | No-CHDD samples DNA (ng/μL) |
| --- | --- | --- | --- | --- | --- | --- | --- |
| BCN01 | ERR13759391 | Not detected | Negative | N/A | N/A | 1.6 | 733 |
| BCN02 | ERR13759392 | Not detected | Negative | N/A | N/A | 2.8 | 767 |
| BCN03 | ERR13759393 | Not detected | Negative | N/A | N/A | 5.5 | 760 |
| BCP01 | ERR13759394 | *S. pyogenes* | Positive | 38.5 | Viable | 209 | 490 |
| BCP02 | ERR13759395 | *S. aureus* | Positive | 32.8 | Viable | 201 | 920 |
| BCP03 | ERR13759396 | *E. coli* | Positive | 50.8 | Viable | 350 | 940 |
| BCP04 | ERR13759397 | *S. pyogenes* | Positive | 38.5 | Viable | 291 | Not tested |
| BCP05 | ERR13759398 | *S. epidermidis* | Positive | 33.0 | Viable | 950 | Not tested |
| BCP06 | ERR13759399 | *S. epidermidis* | Positive | 33.0 | Viable | 620 | Not tested |
| BCP07 | ERR13759400 | *B. luteolum* | Positive | 67.8 | Viable | 17.4 | Not tested |
| BCP08 | ERR13759401 | *M. luteus* | Positive | 74.0 | Not viable | 9.44 | Not tested |
| BCP09 | ERR13759402 | *P. mirabilis* | Positive | 38.8 | Viable | 235 | Not tested |
| BCP10 | ERR13759403 | *S. agalactiae* | Positive | 35.5 | Viable | 77.8 | Not tested |
| BCP11 | ERR13759404 | *E. coli* | Positive | 50.8 | Viable | 1480 | Not tested |
| BCP12 | ERR13759405 | *E. coli* | Positive | 50.8 | Viable | 342 | Not tested |
| BCP13 | ERR13759406 | *K. michiganensis* | Positive | 55.5 | Viable | 1390 | Not tested |
| BCP14 | ERR13759407 | *E. coli* | Positive | 50.8 | Viable | 843 | Not tested |
| BCP15 | ERR13759408 | *K. variicola* | Positive | 57.3 | Viable | 1340 | Not tested |
| BCP16 | ERR13759409 | *S. agalactiae* | Positive | 35.5 | Viable | 325 | Not tested |
| BCP17 | ERR13759410 | *L. adecarboxylata* | Positive | 52.55 | Viable | 111 | Not tested |
| BCP18 | ERR13759411 | *E. coli* | Positive | 50.8 | Viable | 1170 | Not tested |
| BCP19 | ERR13759412 | *S. aureus* | Positive | 32.8 | Viable | 326 | Not tested |
| BCP20 | ERR13759413 | *S. maltophilia* | Positive | 66.7 | Viable | 18.1 | Not tested |
| BCP21 | ERR13759414 | *S. epidermidis* | Positive | 33.0 | Viable | 440 | Not tested |
| BCP22 | ERR13759415 | *E. faecalis* | Positive | 37.5 | Viable | 432 | Not tested |
| BCP23 | ERR13759416 | *K. pneumoniae* | Positive | 57.3 | Viable | 680 | Not tested |
| BCP24 | ERR13759417 | *P. mirabilis* | Positive | 38.7 | Viable | 860 | Not tested |
| BCP25 | ERR13759418 | *E. coli* | Positive | 50.8 | Viable | 1033 | Not tested |
| BCP26 | ERR13759419 | *S. gallolyticus* | Positive | 37.69 | Viable | 297 | Not tested |
| BCP27 | ERR13759420 | *S. aureus* | Positive | 32.8 | Viable | 228 | Not tested |
| BCP28 | ERR13759421 | *A. xylosoxidans* | Positive | 64.0 | Viable | 84 | Not tested |
| BCP29 | ERR13759422 | *S. epidermidis* | Positive | 33.0 | Viable | 406 | Not tested |
| BCP30 | ERR13759423 | *C. jejuni* | Positive | 30.3 | Viable | 348 | Not tested |

**Tab. S2:** Double stranded DNA concentration of three 8-hour culture enriched sterile blood (HB1 to HB3) versus ten spiked blood (1-10 CFU) samples following 60 minutes of multiple displacement amplification with REPLI-g kit with and without M-15 chemical host DNA depletion (CHDD) measured by Qubit. Viability of bacteria following M-15 CHDD was confirmed by plating the supernatant on nutrient and blood agars.

| Sample | Organism | Culture | GC content (%) | Viability after M-15 CHDD | CHDD samples DNA (ng/μL) | No-CHDD samples DNA (ng/μL) |
| --- | --- | --- | --- | --- | --- | --- |
| HB1 | Not detected | Negative | NA | N/A | 4.2 | 652 |
| HB2 | Not detected | Negative | NA | N/A | 8.1 | 487 |
| HB3 | Not detected | Negative | NA | N/A | 4.18 | 513 |
| RESB1 | *S. aureus* | Positive | 32.8 | Viable | 131 | 510 |
| RESB2 | *E. faecium* | Positive | 37.8 | Viable | 122 | 611 |
| RESB3 | *S. pyogenes* | Positive | 38.5 | Viable | 140 | 377 |
| RESB4 | *S. agalactiae* | Positive | 35.6 | Viable | 177 | 647 |
| RESB5 | *S. pneumoniae* | Positive | 39.7 | Not viable | 83 | 550 |
| RESB6 | *P. aeruginosa* | Positive | 66.6 | Viable | 43 | 449 |
| RESB7 | *K. quasipneumoniae* | Positive | 57.2 | Viable | 213 | 714 |
| RESB8 | *A. baumannii* | Positive | 39 | Viable | 156 | 699 |
| RESB9 | *E. coli* | Positive | 50.8 | Viable | 248 | 892 |
| RESB10 | *P. mirabilis* | Positive | 38.88 | Viable | 233 | 621 |

**Tab. S3:**
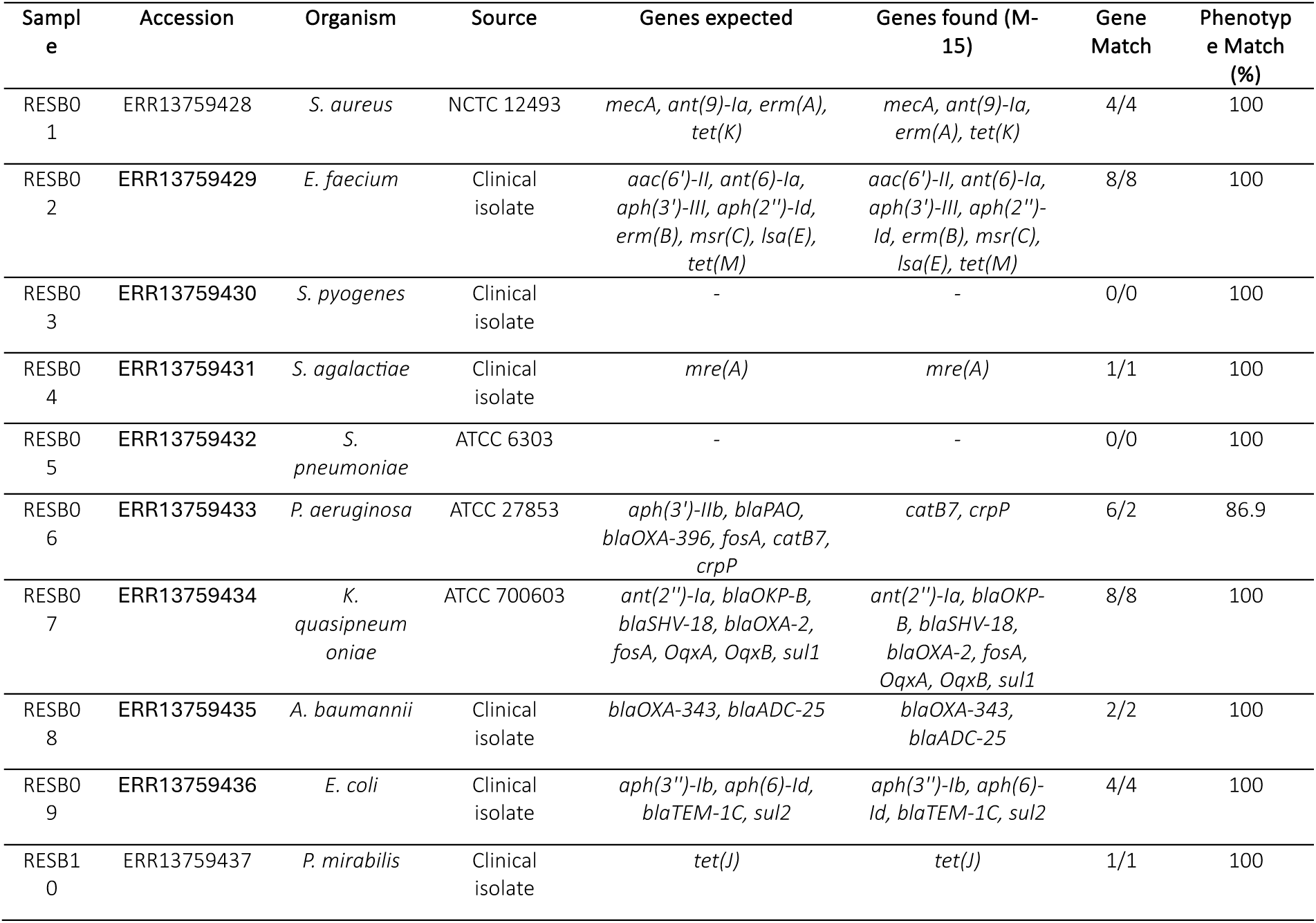
AMR gene profile (genes expected based on whole genome sequencing data) of the 8-hour culture-enriched blood samples chosen for sequencing (n=10). Following M-15 mNGS, sequencing reads were analysed with ResFinder to match and confirm the presence/absence of the same AMR determinants expected from the species initially utilised for spiking. Prediction accuracy of AMR (Match%) was determined comparing the ResFinder predicted phenotypic AST results of M-15 mNGS versus previously sequenced clinical/ATCC strains.

**Tab. S4:**
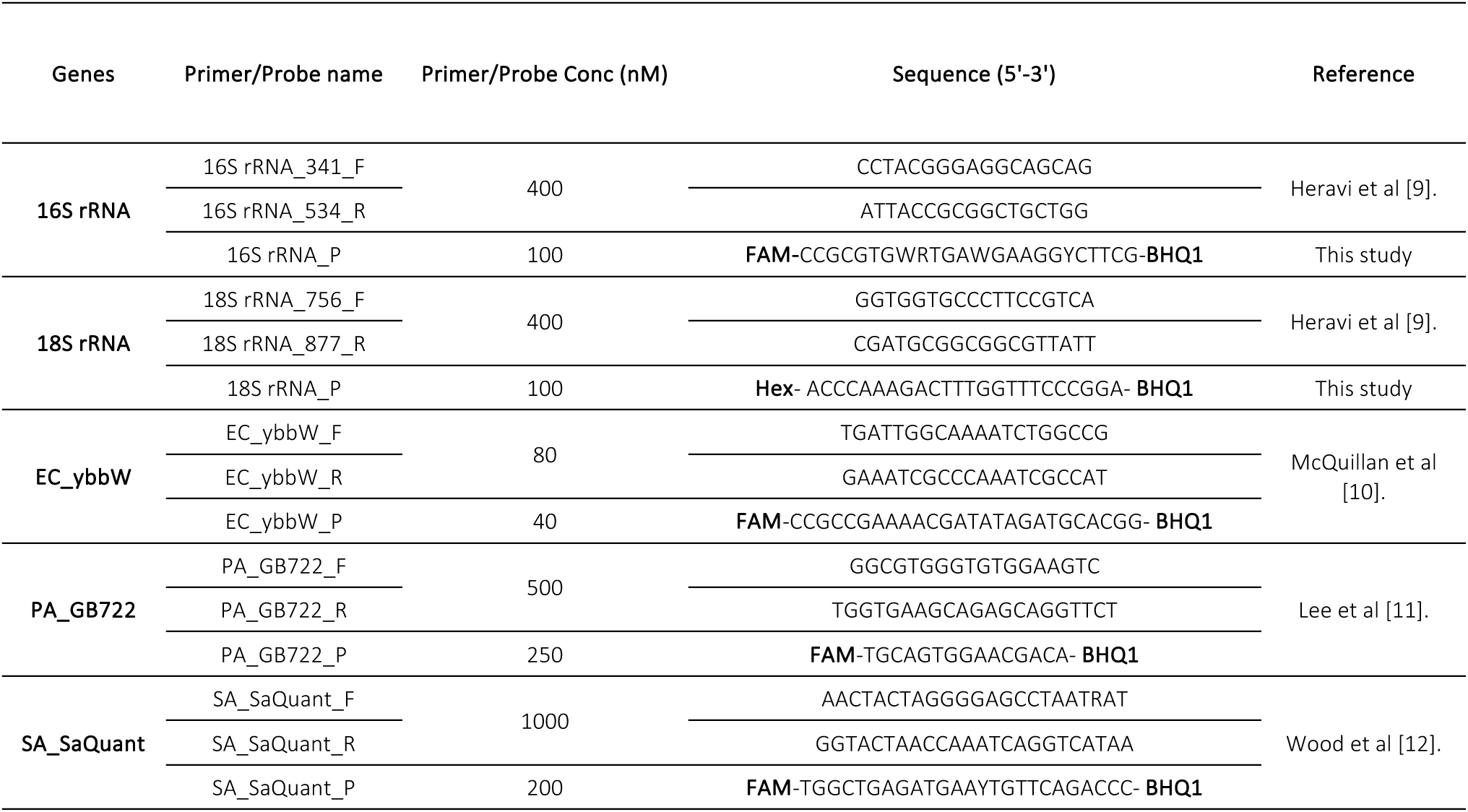
Primer, probe sequences and their reaction concentrations used in this study for detecting universal 16S (bacteria) and 18S Ribosomal ribonucleic acid-rRNA (host) genes and species-specific genes to identify three bacterial species, (*Escherichia coli* (EC), *Pseudomonas aeruginosa* (PA), and *Staphylococcus aureus* (SA)) used for initial spiking and benchmarking experiments.

## Supplement Protocol: M-15 mNGS

### Consumables

Supplement Protocol: M-15 mNGS Consumables
| Name | Concentration/units | Commercial source | Comments |
| --- | --- | --- | --- |
| Saponin | Final concentration<br>2.25% | Tokyo chemical<br>industry, Cat: S0019 | To make a 4.5% or<br>2.25% w/v solution,<br>dissolve 4.5/2.25<br>grams of Saponin into<br>50 ml sterile nuclease<br>free H<sub>2</sub>O. Mix<br>thoroughly until<br>dissolved. Adjust the<br>volume to 100ml.<br>Sterilize using 0.2 µm<br>filter, and store at 4<br>°C for up to 3 weeks |
| NaCl | 5M | Promega, Cat:<br>V4221 | NA |
| DNase I | 300 units/µL | Invitrogen, Cat:<br>18047019 | NA |
| DNase I buffer | 10x solution | NEB, Cat:<br>B0303SVIAL | 10x buffer = 10 mM<br>Tris HCl, 2.5 mM<br>MgCl<sub>2</sub>, 0.5 mM CaCl<sub>2</sub> |
| Trypsin-EDTA | 0.05%, phenol red | ThermoFisher Cat:<br>25300054 | NA |
| Molecular grade<br>H<sub>2</sub>O | NA | Qiagen, Cat: 129114 | NA |
| DPBS | 1x solution | ThermoFisher Cat:<br>14190144 | Without calcium,<br>magnesium, or<br>phenol |
| T7 Endonuclease I | NA | NEB, Cat: M0302S or<br>M0302L | NA |
| DNA binding<br>beads | NA | Aline biosciences,<br>Cat: C-1003-50 | NA |
| Molecular grade<br>ethanol | NA | Sigmaaldrich, Cat:<br>51976-500ML-F | NA |
| REPLI-g Single Cell Kit | NA | Qiagen, Cat: 150343 or 150345 | NA |
| Qubit dsDNA Be assay Kit | NA | ThermoFisher, Cat: Q32853 | NA |
| Qubit assay tubes | NA | ThermoFisher, Cat: Q32856 | NA |
| Rapid Barcoding Kit | NA | ONT, Cat: SQK-RBK004 | Required for the rapid sequencing protocol |
| Long Fragment Buffer Expansion | NA | ONT, Cat: EXP-LFB001 | Required for the rapid sequencing protocol |
| Ligation Sequencing Kit | NA | ONT, Cat: SQK-LSK109 | Required for the high yield protocol |
| Native Barcoding Expansion Kit | NA | ONT, Cat: EXP-NBD196 | Required for the high yield protocol |
| NEBNext Ultra II End Repair/dA-Tailing Module | NA | NEB, Cat: E7546L | Required for the high yield protocol |
| Blunt/TA Ligase Master Mix | NA | NEB, Cat: M0367L | Required for the high yield protocol |
| NEBNext Quick Ligation Module | NA | NEB, Cat: E6056L | Required for the high yield protocol |
| Flow Cell Priming Kit | NA | ONT, Cat: EXP-FLP002 | Required for both protocols |
| Flow Cell Wash Kit | NA | ONT, Cat: EXP-WSH004 | Required for both protocols |
| ONT Flow Cell R9.4.1 | NA | ONT, Cat: FLO-MIN106 or FLO-FLG001 | Required for both protocols |
| Magnetic rack/plate holder | NA | ThermoFisher, Cat: 12331D and 12321D | NA |
| PCR Tube Strip with Attached Flat Caps | NA | ThermoFisher, Cat: AB2000 | NA |
| 1.5 ml DNA LoBind tubes | NA | Eppendorf, Cat: 0030108051-250EA | Use LoBind tubes |
| 2 ml round bottom tubes | NA | Starlab, Cat: S1620-2700-C or Eppendorf, Cat: 0030120094 | Avoid using LoBind tubes |
| Pipettes and pipette tips P2, P10, P20, P100, P200, P1000, Multichannel | NA | Any | NA |
| Centrifuge /Microplate centrifuge | NA | Any | NA |
| Vortex mixer | NA | Any | NA |
| Thermal cycler | NA | Any | NA |
| Heating block for 1.5/2 mL tubes | NA | Any | NA |
| Sequencing device | NA | Flongle, MinION Mk1B, GridION | NA |
| Device for live basecalling | NA | GridION or A powerful laptop | A laptop with minimum 8 Gb GPU (RTX 3060 or higher), 16 GB Ram, 1 Tb M.2 NVMe SSD |

**Disclaimer:** Some parts of the following protocol has been copied directly from the manufacturer's official website or from the kit manual. Therefore, all the proprietary information mentioned below remains the sole property of the kits manufacturer.

### Before sample preparation

It is important to divide all reagents into smaller aliquots to avoid multiple freeze-thaw cycles and reduce the likelihood of cross-contamination. Warm up 0.05% trypsin EDTA at 37°C. Prepare 1x DNase I buffer and store in the fridge before use. Make a fresh batch of REPLI-g Master Mix following manufacturer’s instructions. Take out aliquots of Buffer D2, Stop Solution, and any other reagents from the freezer, and store in the refrigerator or on ice until needed. Thaw all the reagents at room temperature or on ice as per manufacturers recommendation.

This protocol can be used on BD BACTEC (Becton, Dickinson and Company, NJ, USA) or any blood culture (FPBC) samples flagged positive with an automated culture system. Protocol M-15 mNGS can also be applied to rapid culture enriched blood samples (REBC; for example, 1-10 mL patient blood samples incubated for 8 hours in BD BACTEC media bottles), once the enriched sampled have at least 10^3^ CFU/mL bacterial concentration. FPBC samples can be used directly for CHDD; however, REBC samples need to be pre-processed to concentrate the sample before continuing with this protocol. It is crucial to use fresh samples for CHDD, as frozen or improperly stored samples can lead to the lysis of bacterial cells. This may lead to a considerable loss of bacterial DNA during CHDD.

### Pre-processing REBC samples

Transfer 1.5 mL REBC samples aseptically to a 2 mL round bottom microcentrifuge tube and spin at 8,000xg for 5 minutes. Slowly aspirate the supernatant without disturbing the pellet and gently resuspend the pellet in 800 μL 2.25% saponin. Proceed to the step 2 of CHDD protocol below. Note* It is important to use round/wide bottom 2 mL microcentrifuge tubes to minimize bacterial cell loss while aspirating the supernatant during the initial washing steps.

### Host depletion

1. Add 400 μL FPBC sample to 400 μL 4.5% saponin (final saponin concentration will be 2.25% after mixing) in a 2 mL wide bottom sterile microcentrifuge tube and gently pipette mix for at least 4 to 5 times.
2. Add 1 μL DNase I (300 units/μL) to the REBC/FPBC sample, pulse vortex at low speed for 5 seconds and incubate the sample at 25°C in a heating block for 10 minutes at 800 rpm. Note* Adding DNase I decreases the viscosity of the sample by degrading a substantial amount of DNA (desirably from the host) after selective host cell lysis. The DNA released from the host cells can be extremely sticky, and often adhere to pipette tips or microfuge tubes. As a result, there’s a risk of losing desired/targeted bacterial cells because they might adhere to the sticky host DNA and get affixed to the pipette tips and/or microcentrifuge tube.
3. Perform a quick spin and add 800 μL molecular grade water to the sample. Pipette mix 4 to 5 times and incubate for 1 minute at room temperature.
4. Add 44 μL 5 M molecular grade NaCl (final concentration is 137 mM; same as PBS) to the sample, invert the tubes 3 to 4 times to mix and centrifuge at 10,000x g for 5 minutes.
5. Carefully remove the supernatant without touching or disturbing the pellet and resuspend in 1 mL molecular grade 1x Dulbecco’s Phosphate Buffered Saline (DPBS, Without calcium, magnesium, or phenol).
6. Centrifuge the sample at 10,000x g for 3 minutes and carefully remove the supernatant without touching or disturbing the pellet.
7. Resuspend the pellet in 398 μL 1x DNase I buffer, add 2 μL DNase I (300 units/μL) to the sample and incubate at 37°C with shaking at 800 RPM for 15 minutes.
8. Following incubation, add 1 mL 0.05% trypsin EDTA directly to the sample, pipette mix at least 8 to 10 times and incubate at room temperature for 1 minute. Note* Some bacterial cells may form aggregates in the sample, so adding 0.05% trypsin EDTA can separate the cells and ensure an even distribution for subsequent procedures. The trypsinisation step does not affect bacterial viability and/or cell wall integrity.
9. Repeat step 6 and resuspend the pellet in 1 mL 1x DPBS. Note* This is the end of host depletion with M-15. After this, 4 μL of host depleted bacterial cell material in DPBS from step 9 can be used directly for alkaline lysis and whole genome amplification (WGA; protocol 1) using the REPLI-g Single Cell Kit. **DNA extraction and Whole Genome Amplification**
10. Prepare sufficient Buffer D2 in a PCR tube for the total number of 12 reactions.

11. Place 4 µL cell material into a PCR tube containing 3 µL buffer D2. Mix by flicking the tube and centrifuge briefly.
12. Incubate at 65°C for 10 min with heated lid on and then add 3 µL Stop Solution. Mix by flicking the tube and centrifuge briefly. Prepare a master mix according to the instruction below.

| Reagent | Volume (µL) |
| --- | --- |
| DTT, 1M | 3 |
| Buffer DLB | 33 |
| Total | 36 |

Note\* Buffer D2 should not be stored longer than 2 months at -20 °C.

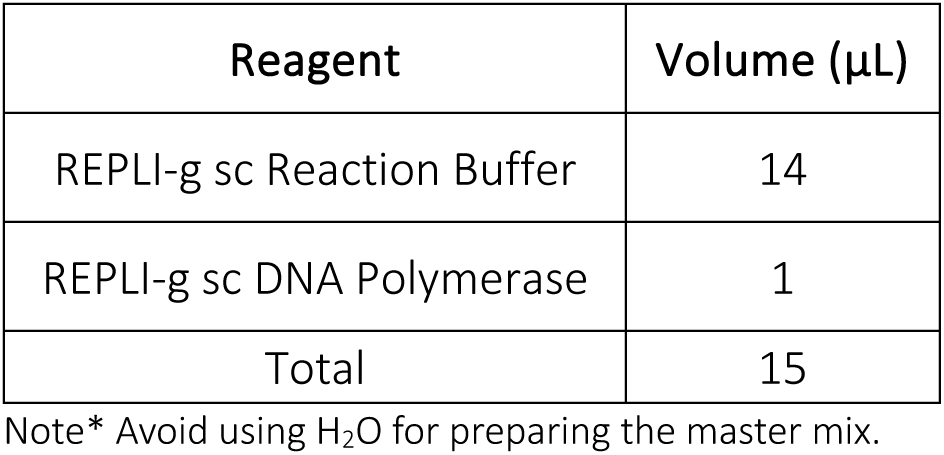
13. For each reaction, add 5 µL DNA from step 12 and incubate at 30°C for 1 hour with heated lid off.
14. Inactivate REPLI-g sc DNA Polymerase by heating the sample for 3 min at 65°C with heated lid on. Note* After this step amplified host depleted DNA can be diluted 5-fold in molecular grade H2O, measured with Qubit Br assay and used directly for debranching and subsequently sequencing library preparation. **Debranching and bead cleanup**
15. In a clean 0.2 mL PCR tube, mix the reagents for debranching in the following order.

16. Mix the reagents by flicking the tube and centrifuge briefly. Incubate the reaction for 10 minutes at 37°C.
17. Resuspend the DNA binding beads (e.g., AMPure XP or Aline biosciences) by vortexing.
18. Add 15 µL of the bead suspension (0.5x) to the DNA sample and mix by pipetting.
19. Incubate for 5 minutes at room temperature.
20. Spin down and pellet the sample on a magnetic rack/plate until supernatant is clear and colourless. Keep the tube on the magnet, and pipette off the supernatant.
21. Keeping the tube on the magnet, wash the beads with 200 µL of freshly prepared 70% ethanol without touching/disturbing the pellet. Remove the ethanol carefully using a pipette and discard.
22. Repeat the previous ethanol washing step.
23. Spin down and put the tube back on the magnetic rack/plate. Pipette off any residual ethanol. Open the lid and allow the tubes to dry for ∼30 seconds, but do not dry the pellet to the point of cracking.
24. Remove the tube from the magnetic rack/plate and resuspend the pellet in 15 µL nuclease-free water. Incubate for 2 min at room temperature.
25. Pellet the beads on a magnet until the eluate is clear and colourless.
26. Remove and retain 12 µL of eluate into a clean 0.2 mL PCR tube. Note* This is the end of debranching and DNA cleanup. After this step, cleaned and debranched DNA products can be used for library preparation with Rapid Barcoding (rapid but low yield) or Ligation sequencing kit (highest yield).

| Reagent | Volume (μL) |
| --- | --- |
| 600 ng of amplified DNA | To make 30 μL |
| NEBuffer 2 | 3 |
| T7 Endonuclease I | 1.5 |
| Nuclease free H<sub>2</sub>O | 25.5- To make 30 μL |
| Total | 30 |

### Rapid barcoding (low-medium yield protocol) Barcoding

1. Combine the following components per PCR tube:

2. Mix the reagents by flicking the tube and spin briefly.
3. Incubate the samples at 30°C in a thermal cycler for 1 minute and 80°C for 1 minutes with heated lid off.
4. Spin and pool all the rapid barcoded DNA in 1.5 mL Eppendorf DNA LoBind tube to make 1,200 ng in a volume of 50 µL. Example, for 6 samples, take 200 ng (200 x 6 = 1,200 ng) DNA per sample.
5. Resuspend the DNA binding beads (e.g., AMPure XP or Aline biosciences) by vortexing.
6. Add 25 µL (0.5x) of resuspended beads to the sample and mix by flicking the tube multiple times.
7. Incubate the tube for 5 minutes at room temperature.
8. Spin down the sample and pellet on a magnetic rack/plate until the supernatant is clear and colourless. Keep the tube on the magnet, and carefully pipette off the supernatant.
9. Wash the beads with 250 μl long Fragment Buffer (LFB) without touching/disturbing the pellet. Remove LFB carefully using a pipette and discard. Note* Flick the beads to resuspend, spin and put back on magnet and remove supernatant using a pipette. Long Fragment Buffer Expansion (EXP-LFB001) can be purchased separately from ONT.
10. Repeat the previous LFB washing step.
11. Spin down and place the tube back on the magnet. Pipette off any residual supernatant. Allow to dry for ∼30 seconds, but do not dry the pellet to the point of cracking.
12. Remove the tube from the magnetic rack/plate and resuspend the pellet in 22 µL Elution Buffer (EB). Spin down briefly and incubate for 2 minutes at room temperature.
13. Pellet the beads on a magnetic rack/plate until the eluate is clear and colourless, for at least 1 minute.
14. Remove and retain 20 µL of eluate containing the DNA library into a clean 1.5 mL Eppendorf DNA LoBind tube. **Rapid adapter ligation and loading**
15. Quantify 1 µL rapid barcoded and cleaned DNA using a Qubit fluorometer.
16. Add 1 µL rapid adaptor (RAP) to 10 µL of barcoded DNA in EB in a PCR tube, flick multiple times to mix, spin briefly and incubate for 10 minutes at 20°C with the heated lid off.
17. Load 100 ng (51.36 fmol; considering 3kb product size) of final prepared library onto a flow cell following manufacturers recommendation.

| Reagent | Volume (µL) |
| --- | --- |
| Debranched, cleaned<br>DNA (400 ng) | 7.5 |
| Fragmentation Mix<br>(RB01-12) | 2.5 |
| Total | 10 |

## Ligation sequencing (high yield protocol)

### End prep

1. Combine the following components per PCR tube:

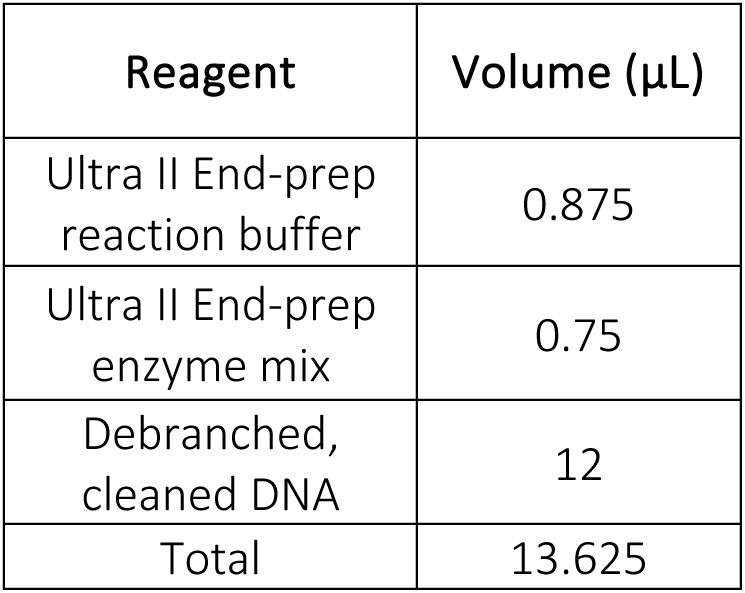
2. Mix gently by flicking the tube, and spin down.
3. Using a thermal cycler, incubate at 20°C for 5 minutes with heated lid off.
4. Incubate at 65°C for 5 minutes with heated lid on. **Native barcode ligation and cleanup**
5. Combine the following components per PCR tube:

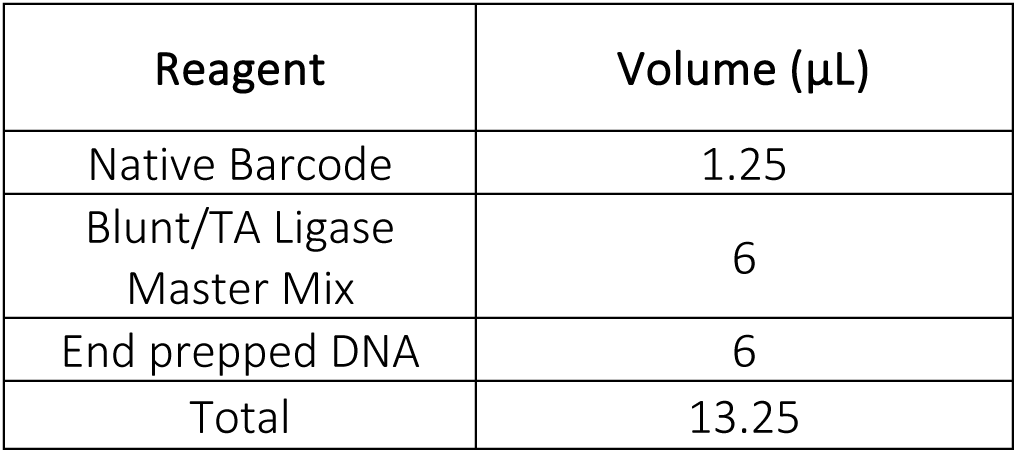
6. Mix gently by flicking the tube, and spin down.
7. Incubate the reaction for 10 minutes at 20°C with heated lid off.
8. Resuspend the AMPure XP beads by vortexing.
9. Add 13.25 µL of resuspended AMPure XP beads to the reaction and mix by pipetting.
10. Incubate for 5 minutes at room temperature.
11. Spin down the sample and pellet on a magnet until supernatant is clear and colourless. Keep the tube on the magnet, and pipette off the supernatant.
12. Keep the tube on the magnet and wash the beads with 200 µL of freshly prepared 70% ethanol without disturbing the pellet. Remove the ethanol using a pipette and discard.
13. Repeat the previous step.
14. Spin down and place the tube back on the magnet. Pipette off any residual supernatant. Allow to dry for ∼30 seconds, but do not dry the pellet to the point of cracking.
15. Remove the tube from the magnetic plate and resuspend pellet in 15 µL nuclease-free water. Incubate for 2 min at room temperature.
16. Pellet the beads on a magnet until the eluate is clear and colourless.
17. Remove and retain 12 µL of eluate into a clean PCR tube. Measure DNA concentration with Qubit and pool all the barcoded DNA in 1.5 mL Eppendorf DNA LoBind tube to make 350 ng in a volume of 30 µL H2O. Example, for 6 samples, take 58.33 ng (58.33 x 6 = 350 ng) DNA per sample. **Adapter ligation and cleanup and loading**
18. Combine the following components:

| Reagent | Volume (μL) |
| --- | --- |
| Native Barcode | 1.25 |
| Blunt/TA Ligase Master Mix | 6 |
| End prepped DNA | 6 |
| Total | 13.25 |

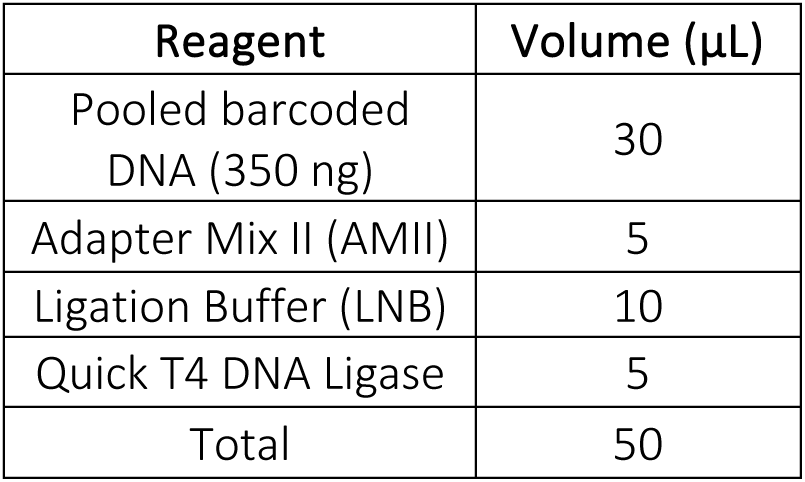
19. Ensure the components are thoroughly mixed by pipetting, and spin down.
20. Incubate the reaction for 15 minutes at 20°C.
21. Resuspend the AMPure XP beads by vortexing.
22. Add 25 µL (0.5x) of resuspended AMPure XP beads to the reaction and mix by pipetting.
23. Incubate for 5 minutes at room temperature.
24. Spin down the sample and pellet on a magnet until supernatant is clear and colourless. Keep the tube on the magnet, and pipette off the supernatant.
25. Wash the beads by adding 250 μl long Fragment Buffer (LFB). Note* Do not flick the beads to resuspend. Just gently wash and remove the supernatant using a pipette and discard.
26. Repeat the previous step.
27. Spin down and place the tube back on the magnet. Pipette off any residual supernatant. Allow to dry for ∼30 seconds, but do not dry the pellet to the point of cracking.
28. Remove the tube from the magnetic rack and resuspend the pellet in 17 µL Elution Buffer (EB). Spin down and incubate for 2 minutes at room temperature.
29. Pellet the beads on a magnet until the eluate is clear and colourless, for at least 1 minute.
30. Remove and retain 15 µL of eluate containing the DNA library into a clean 1.5 mL Eppendorf DNA LoBind tube.
31. Quantify 1 µL of adapter ligated and barcoded DNA using a Qubit fluorometer.
32. Load 100 ng (51.36 fmol; considering 3kb) of final prepared library onto a flow cell following manufacturers recommendation. Note* The typical library size after this stage is around 3 kb. However, users can verify this by running the library on a gel or TapeStation for a few batches. Once the average expected library size is determined (sample or site-wise), there will be no need to repeat this for every batch of samples.

For loading sequencing library or washing flow cells, follow the manufacturers recommended protocols.

